# Burden of fatigue in compensated chronic liver disease: findings from the multinational a:GAP Study

**DOI:** 10.64898/2026.08.28.26361618

**Authors:** Gourdas Choudhuri, Gamar Akhundova-Unadkat, Nivantha Naidoo, Mauricio Morales-Castillo, Xavier Guillaume, Ruben G. Duijnhoven, Azadeh Safaei, Mark G. Swain

## Abstract

**Background & Aims:** Fatigue is a central symptom of chronic liver disease (CLD), substantially impacting health-related quality of life (HRQoL). This study aimed to further understand CLD symptomatology, including fatigue, and its impact on HRQoL from a patient perspective.

**Methods:** **A**bbott **G**lobal **A**ssessment of **P**atients unmet needs (aGAP) was a multinational, cross-sectional survey in adults with compensated CLD in China, India and Mexico, conducted between July and November 2024. Adult participants who self-reported that they had physician-diagnosed CLD and were experiencing fatigue completed a quantitative survey to assess symptom burden and included three HRQoL patient-reported outcome (PRO) questionnaires (Patient-Reported Outcomes Measurement Information System [PROMIS]-29+2, Work Productivity and Activity Impairment – Specific Health Problem version 2.0 [WPAI: SHP], Multidimensional Fatigue Inventory [MFI]).

**Results:** Overall, 505 participants (China: 200; Mexico: 105; India: 200) completed the study. Participants reported that their CLD-related fatigue sometimes, often or always affected their self-esteem/confidence (45.1%) and ability to maintain or acquire new employment (38.6%). Most participants reported moderate (51.3%) or serious (26.9%) fatigue, with 33.5% experiencing fatigue every day or almost every day. Many participants felt their social life was negatively impacted by their fatigue (47.3%) and that there were related financial difficulties (53.9%). Use of validated PRO tools demonstrated severe fatigue (MFI: overall mean [SD] 13.9 [3.4] general fatigue and 13.4 [3.6] physical fatigue) as well as substantial levels of work and activity impairment (WPAI: SHP overall mean [SD] 53.0 [26.4]) and high levels of anxiety, pain interference, depression and sleep interference (PROMIS T-scores ≥54).

**Conclusions:** Fatigue has a substantial impact on HRQoL among adults with CLD across several countries, highlighting a global unmet need for targeted interventions to effectively identify and manage the condition.

**Highlights:**

- In this multinational survey of 505 adults with compensated CLD, one-third experienced fatigue daily or almost daily, with substantial impact on social function, employment and HRQoL.
- Fatigue co-occurred with elevated anxiety, depression, pain interference and sleep disturbance (PROMIS T-scores ≥54), along with pronounced work and activity impairment (WPAI:SHP 53.0).
- Cross-country (India, China, Mexico) differences in fatigue perception and support largely reflected sociodemographic and healthcare-system factors rather than biological variation in CLD.
- These findings demonstrate a global unmet need for systematic assessment and targeted management of fatigue in routine CLD care.

## Introduction

Chronic liver disease (CLD) is a leading cause of morbidity and mortality worldwide, accounting for 1.32 million deaths in 2017^1,2^ The most common causes of CLD have shifted in the last 30 years, with viral hepatitis declining in prominence and obesity and alcohol consumption becoming key CLD risk factors.^2^ These patterns vary geographically and are shaped by country-specific health policy and practice for the prevention and management of the different CLD causes.^3^

Patients with CLD experience a broad range of symptoms depending on disease stage and the underlying disease process.^4^ Fatigue, reported to impact between 50 and 85% of patients with CLD,^5^ is composed of peripheral and central causes, and is often associated with behavioral changes such as depression and anxiety.^5–7^ Central fatigue can be characterized by lack of self-motivation, manifesting in both physical and mental symptoms, while peripheral fatigue generally presents as neuromuscular dysfunction and muscle weakness.^8^ Importantly, fatigue has substantial deleterious effects on health-related quality of life (HRQoL) and physical and social activities, seriously impacting patients’ day-to-day lives.^7^ As an “invisible” symptom, fatigue can be difficult to identify, characterize and differentiate from other similar symptoms such as somnolence, weakness and lethargy.^8^ As such, it is often missed by healthcare providers. However, its debilitating nature is clear, therefore better understanding and management of fatigue in CLD is greatly needed.

Patient-reported outcomes are increasingly important components of clinical research, providing valuable insights into the patient experience.^9^ In CLD, these measures have improved the reliability and sensitivity with which fatigue severity and impact can be described.^8^ Precise and accurate assessment of fatigue should improve management leading to better patient outcomes. The aim of this study was to further understand CLD symptomatology, including fatigue, and its impact on HRQoL through administration of a patient survey and relevant patient-reported outcome tools.

## Methods

### Study design

aGAP (**A**bbott **G**lobal **A**ssessment of **P**atients unmet needs) was a multinational, web-based, cross-sectional survey study in adults with CLD in China, India and Mexico, conducted between July and November 2024. Participants completed a survey to assess symptom burden and the perceived impact of fatigue. The survey instrument, a quantitative questionnaire, was developed based on an adapted targeted literature review^10^ conducted on Ovid to identify articles and conference abstracts published between 2013 and 2023 covering observational studies reporting symptomatology and HRQoL in adults diagnosed with CLD **(Supplementary Table 1**). Relevant studies were included for data extraction and thematic analysis to inform question and response formulation. The survey was then tested in qualitative interviews with a small sample of patients with CLD in each country to evaluate appropriateness, clarity and coherency. During the interviews, consistency of interpretation across respondents was assessed as were survey conciseness and manageability among respondents. For each question of the survey instrument, the moderator was asked to probe on whether the question was easy or difficult to answer and, if relevant, what the respondent would change. Explanations of key terms were added where necessary, and it was checked that no item(s)/question(s) of importance were missing. CLD status was established by self report to screener items confirming a prior physician diagnosis and etiology (e.g., viral hepatitis, metabolic steatotic liver disease, alcohol related disease, autoimmune cholestatic disease). Compensated status was determined by negative responses to prespecified decompensation prompts (history of ascites, variceal bleeding, hepatic encephalopathy, or jaundice attributed to decompensation) and by excluding participants who endorsed any such events. The study was approved by country-specific Independent Ethics Committees (i.e., India: Royal Pune Independent Ethics committee; China: Shanghai Clinical Research Ethics Committee; Mexico: CEISH-USFQ University of San Francisco Ethics Committee) and conducted according to the principles of the Declaration of Helsinki, Good Pharmacoepidemiology Practices, and Oracle Health and Life Sciences standard operating procedures. Written e consent was obtained from all participants before completing any study procedures.

### Participants

The study team partnered with an international data collection agency, with recruitment conducted by local recruiters in each country who specialized in healthcare studies and had extensive experience in communicating and engaging with patients. A multimodal approach was used that included physicians’ referral (in all countries), referral through relevant CLD patient associations and support groups (in Mexico and India only), and patient databases (in China and Mexico only). It should be noted that the databases are not specific to CLD; they are databases of respondents to previous research (consumer, market, or observational research) and for whom various profile information has been stored, including some health conditions they may have.

To facilitate physician referral, local recruiters contacted physicians and explained the study specifications, asking them to disseminate the study information to their patients. For these participants, it was specified that the study should not interfere with standard medical care, no additional visits, tests or procedures should be conducted, and no patient data from medical records should be abstracted or communicated by the physician.

To facilitate CLD patient association referral, local recruiters contacted relevant patient associations and support groups and requested that they engage their eligible members, asking them to contact recruiters if interested in participating in the study.

Patient databases were housed by local partners in China (Advanced Healthcare Research Co. Ltd.) and Mexico (Exafield). Patients were recruited to the databases through physician referral, snowball recruitment, conferences (mainly for healthcare professionals) and desk research. Data were quality checked to avoid record duplication and proof of diagnosis/treatment was requested where required for the study. Written or verbal informed consent for inclusion in the database was gathered for all patients, and additional consent was requested for study participation. Consent could be withdrawn at any time.

Eligible participants were adults (≥18 years of age) who self-reported that they had physician-diagnosed CLD and were experiencing fatigue. Participants currently taking any prescribed pharmacologic treatments specifically for fatigue were excluded as were those with decompensated cirrhosis. All participants provided informed consent.

### Study procedures

Eligible participants self-completed an online screener for study participation qualification based on the eligibility criteria. After screener qualification and e-consent provision, all participants completed the main survey. The survey covered sociodemographic data, clinical data, treatments, impact of CLD and fatigue on HRQoL, medical consultations/hospital visits, socio-behavioral characteristics, and general health data. Participants with fully completed surveys were compensated according to fair market value.

Following the main survey, participants completed globally recognized and validated patient-reported outcome measures (PROMs), namely the Patient-Reported Outcomes Measurement Information System (PROMIS)-29+2, the Work Productivity and Activity Impairment – Specific Health Problem (WPAI:SHP), the Multidimensional Fatigue Inventory (MFI) and the Alcohol Use Disorders Identification Test-Concise (AUDIT-C). PROMIS-29+2 is a 29 plus 2 item scale designed to measure eight dimensions of health status (physical function, anxiety, depression, fatigue, sleep disturbance, ability to participate in social roles and activities, pain, cognitive function – abilities) that has been validated for use in patients with CLD.^11^ The WPAI:SHP, validated in chronic conditions such as ulcerative colitis and rheumatoid arthritis,^12, 13^ is a 6-item scale measuring impairments in work productivity and everyday life activities. The MFI, utilized across multiple patient populations,^14^ is a 20-item scale designed to evaluate five dimensions of fatigue: general fatigue, physical fatigue, reduced motivation, reduced activity and mental fatigue. AUDIT-C is a validated 3-question scale (5 items per question) measuring alcohol consumption per week in the past 12 months and alcohol consumption per occasion in a variety of populations.^15^

### Statistical analysis

As this was a descriptive study with no hypothesis testing, there was no formal sample size calculation required, however a sample size of 500 participants (200 from China, 200 from India and 100 from Mexico) was deemed to be large enough to allow robust descriptive subgroup analyses at a global level. All participants meeting the inclusion criteria and quality checks were included in the final analysis. Data were summarized using descriptive statistics, overall and by country.

A pre-specified subgroup analysis was conducted among subgroups with characteristics of interest including CLD duration (<1 year vs 1–<2 years vs 2–<4 years vs ≥4 years), sex (male vs female), age (<30 years vs 30–39 years vs 40–49 years vs ≥50 years), etiological CLD subtype (non-alcoholic fatty liver disease [NAFLD]/ non-alcoholic steatohepatitis [NASH]/ metabolic dysfunction-associated steatotic liver disease [MASLD]/ metabolic dysfunction-associated steatohepatitis [MASH] vs other; alcohol-related liver disease vs other), viral hepatitis (hepatitis vs other), severe fatigue as assessed by MFI score (≤60 vs >60) and exposure to CLD treatment (currently treated vs treated in the past vs never treated). Bivariate comparisons of subpopulations of interest were conducted using traditional parametric independent-sample t-tests or one-way analysis of variance (ANOVA) tests; p-values <0.05 (two-tailed) were considered statistically significant without adjustment for multiplicity and should be interpreted as exploratory. Although the MFI does not have a universally established severity categorization, prior psychometric work has used higher total scores (≈60 or above) to identify individuals with clinically meaningful or high burden fatigue, and this study applied a similar cut point for exploratory subgroup analyses.

## Results

### Study population

Of 968 participants who entered the survey (464 from China, 208 from Mexico, 296 from India), 381 did not meet the eligibility criteria, 50 did not complete the survey and 32 were removed for quality reasons. This left 505 participants (200 from China, 105 from Mexico, 200 from India). Overall, 61.0% of participants from China, 55.5% of participants from India and 18.1% of participants from Mexico were recruited to the survey through physician referral.

### Main questionnaire

Demographic and clinical information from the main questionnaire is shown in **Table 1**, **Figure 1** and **Supplementary Tables 2-6**. The overall population had a mean (standard deviation [SD]) age of 41.3 (12.6) years and 46.7% were female (**Table 1** and **Supplementary Table 2**). A majority of participants reported being educated to college/university level (55.6%), with the lowest proportion in India (43.5%) and the highest proportion in China (67.5%). Around a quarter of participants were educated to high school level (24.0%). Around half of the overall population (47.9%) were employed full-time, but this proportion varied greatly across countries with 71.0% of participants in China working full-time compared to just 29.5% in Mexico and 34.5% in India. The most common insurance types in China and Mexico, respectively, were Urban Employee Medical Insurance (79.0%) and Instituto Mexicano del Seguro Social (45.7%). In India, almost half of participants (43.0%) answered ‘none of the above’ when given public health insurance, individual/family private insurance plans and private insurance cover through parent/guardian employer as health insurance options.

**Figure 1.**
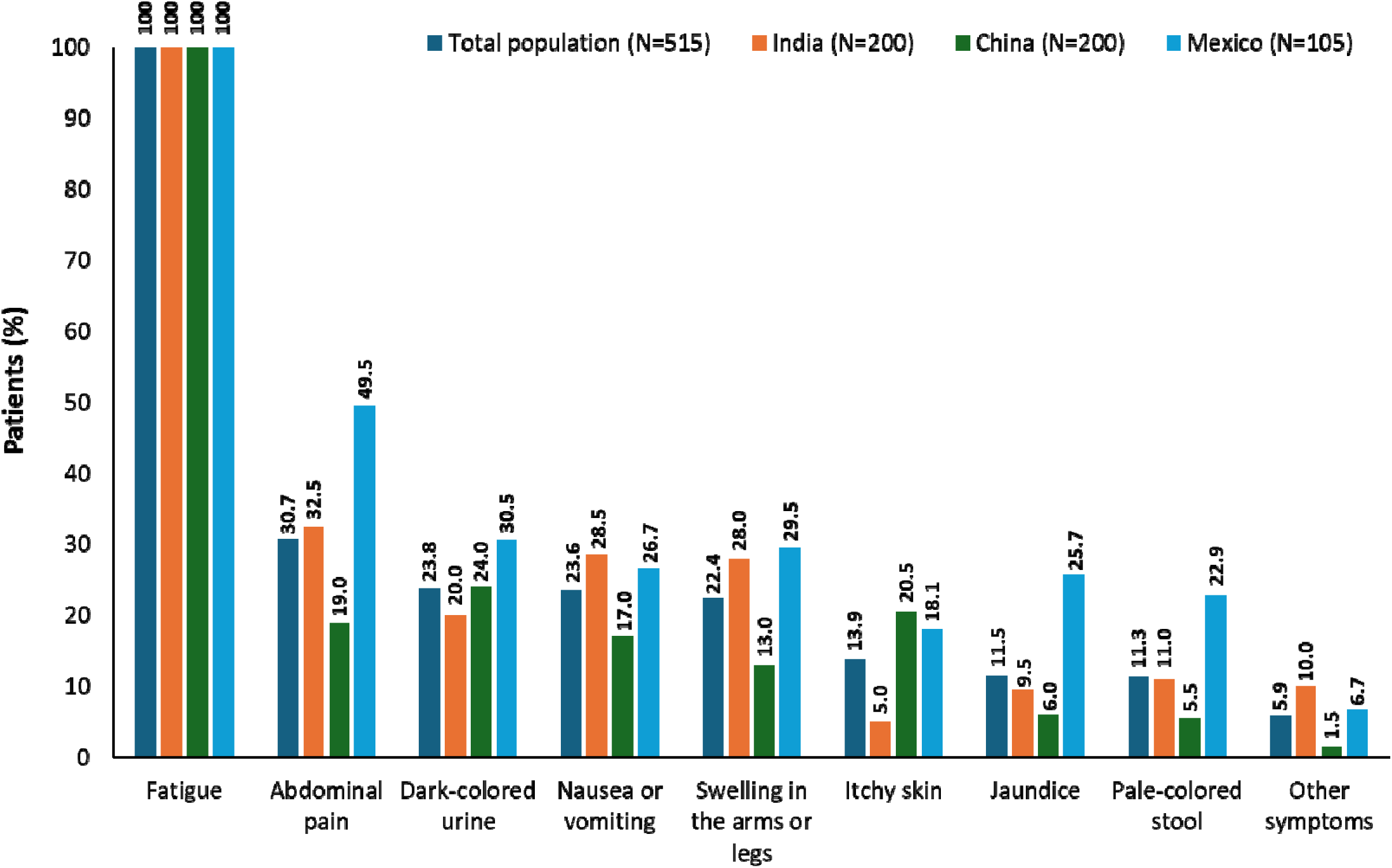
CLD symptoms reported during the main questionnaire CLD, chronic liver disease.

**Table 1.**
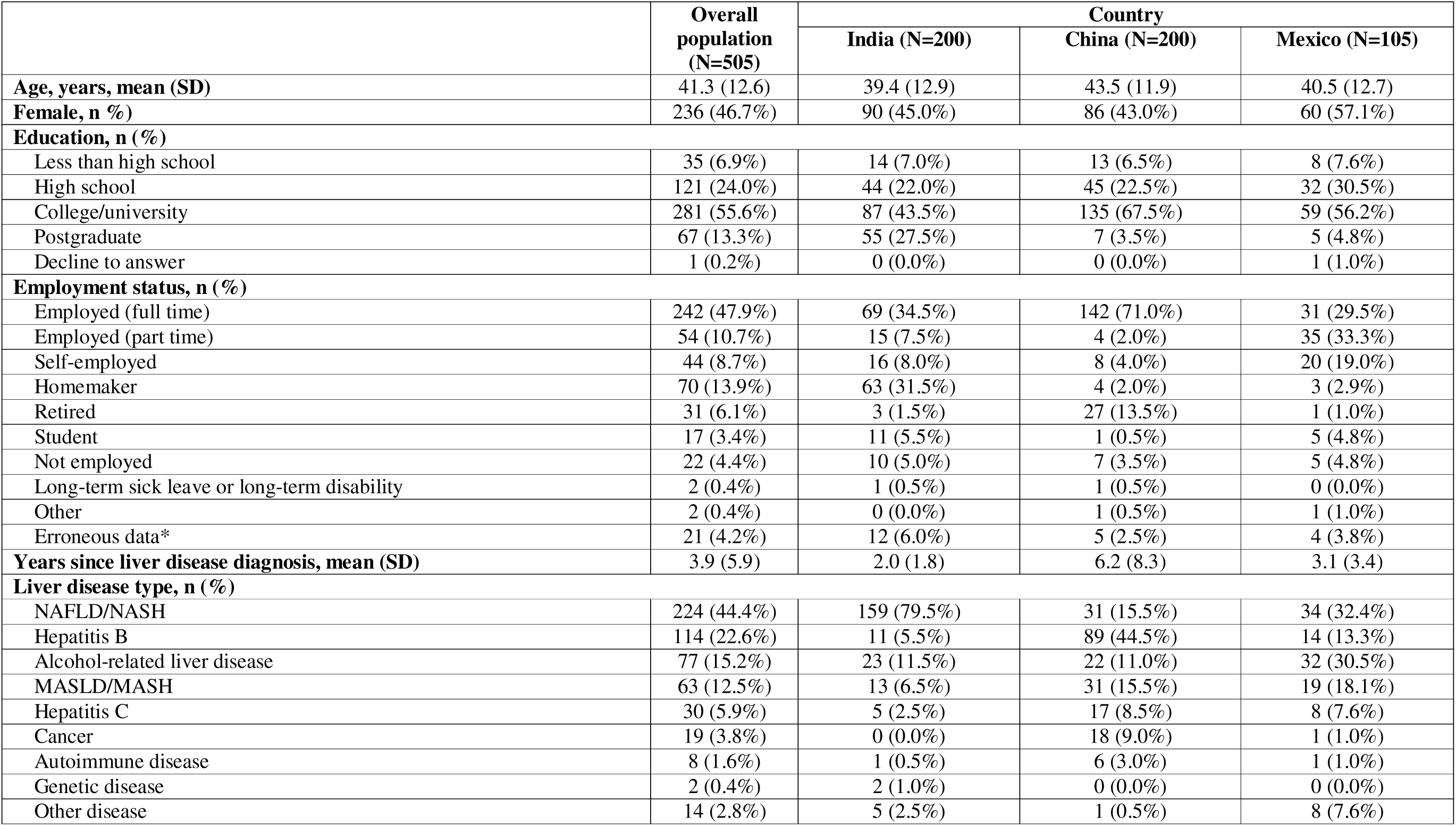
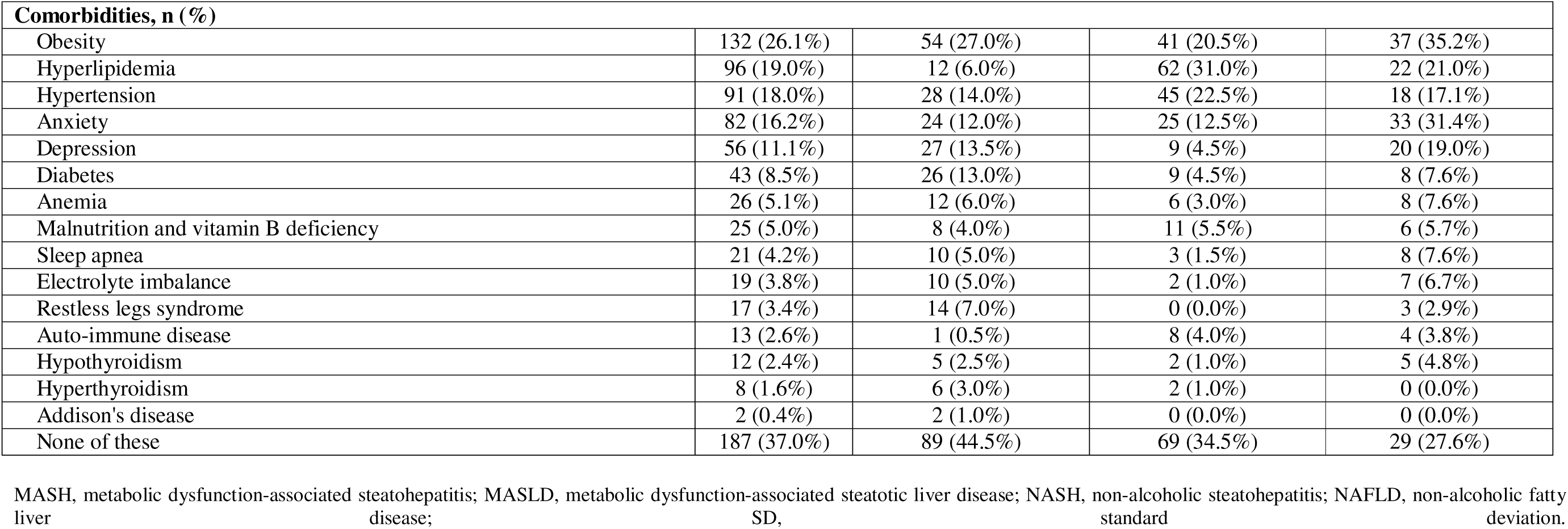
Demographic and clinical characteristics.

|  | Overall population (N=505) | Country |  |  |
| --- | --- | --- | --- | --- |
|  |  | India (N=200) | China (N=200) | Mexico (N=105) |
| **Age, years, mean (SD)** | 41.3 (12.6) | 39.4 (12.9) | 43.5 (11.9) | 40.5 (12.7) |
| **Female, n (%)** | 236 (46.7%) | 90 (45.0%) | 86 (43.0%) | 60 (57.1%) |
| **Education, n (%)** |  |  |  |  |
| Less than high school | 35 (6.9%) | 14 (7.0%) | 13 (6.5%) | 8 (7.6%) |
| High school | 121 (24.0%) | 44 (22.0%) | 45 (22.5%) | 32 (30.5%) |
| College/university | 281 (55.6%) | 87 (43.5%) | 135 (67.5%) | 59 (56.2%) |
| Postgraduate | 67 (13.3%) | 55 (27.5%) | 7 (3.5%) | 5 (4.8%) |
| Decline to answer | 1 (0.2%) | 0 (0.0%) | 0 (0.0%) | 1 (1.0%) |
| **Employment status, n (%)** |  |  |  |  |
| Employed (full time) | 242 (47.9%) | 69 (34.5%) | 142 (71.0%) | 31 (29.5%) |
| Employed (part time) | 54 (10.7%) | 15 (7.5%) | 4 (2.0%) | 35 (33.3%) |
| Self-employed | 44 (8.7%) | 16 (8.0%) | 8 (4.0%) | 20 (19.0%) |
| Homemaker | 70 (13.9%) | 63 (31.5%) | 4 (2.0%) | 3 (2.9%) |
| Retired | 31 (6.1%) | 3 (1.5%) | 27 (13.5%) | 1 (1.0%) |
| Student | 17 (3.4%) | 11 (5.5%) | 1 (0.5%) | 5 (4.8%) |
| Not employed | 22 (4.4%) | 10 (5.0%) | 7 (3.5%) | 5 (4.8%) |
| Long-term sick leave or long-term disability | 2 (0.4%) | 1 (0.5%) | 1 (0.5%) | 0 (0.0%) |
| Other | 2 (0.4%) | 0 (0.0%) | 1 (0.5%) | 1 (1.0%) |
| Erroneous data* | 21 (4.2%) | 12 (6.0%) | 5 (2.5%) | 4 (3.8%) |
| **Years since liver disease diagnosis, mean (SD)** | 3.9 (5.9) | 2.0 (1.8) | 6.2 (8.3) | 3.1 (3.4) |
| **Liver disease type, n (%)** |  |  |  |  |
| NAFLD/NASH | 224 (44.4%) | 159 (79.5%) | 31 (15.5%) | 34 (32.4%) |
| Hepatitis B | 114 (22.6%) | 11 (5.5%) | 89 (44.5%) | 14 (13.3%) |
| Alcohol-related liver disease | 77 (15.2%) | 23 (11.5%) | 22 (11.0%) | 32 (30.5%) |
| MASLD/MASH | 63 (12.5%) | 13 (6.5%) | 31 (15.5%) | 19 (18.1%) |
| Hepatitis C | 30 (5.9%) | 5 (2.5%) | 17 (8.5%) | 8 (7.6%) |
| Cancer | 19 (3.8%) | 0 (0.0%) | 18 (9.0%) | 1 (1.0%) |
| Autoimmune disease | 8 (1.6%) | 1 (0.5%) | 6 (3.0%) | 1 (1.0%) |
| Genetic disease | 2 (0.4%) | 2 (1.0%) | 0 (0.0%) | 0 (0.0%) |
| Other disease | 14 (2.8%) | 5 (2.5%) | 1 (0.5%) | 8 (7.6%) |

| **Comorbidities, n (%)** |  |  |  |  |
| --- | --- | --- | --- | --- |
| Obesity | 132 (26.1%) | 54 (27.0%) | 41 (20.5%) | 37 (35.2%) |
| Hyperlipidemia | 96 (19.0%) | 12 (6.0%) | 62 (31.0%) | 22 (21.0%) |
| Hypertension | 91 (18.0%) | 28 (14.0%) | 45 (22.5%) | 18 (17.1%) |
| Anxiety | 82 (16.2%) | 24 (12.0%) | 25 (12.5%) | 33 (31.4%) |
| Depression | 56 (11.1%) | 27 (13.5%) | 9 (4.5%) | 20 (19.0%) |
| Diabetes | 43 (8.5%) | 26 (13.0%) | 9 (4.5%) | 8 (7.6%) |
| Anemia | 26 (5.1%) | 12 (6.0%) | 6 (3.0%) | 8 (7.6%) |
| Malnutrition and vitamin B deficiency | 25 (5.0%) | 8 (4.0%) | 11 (5.5%) | 6 (5.7%) |
| Sleep apnea | 21 (4.2%) | 10 (5.0%) | 3 (1.5%) | 8 (7.6%) |
| Electrolyte imbalance | 19 (3.8%) | 10 (5.0%) | 2 (1.0%) | 7 (6.7%) |
| Restless legs syndrome | 17 (3.4%) | 14 (7.0%) | 0 (0.0%) | 3 (2.9%) |
| Auto-immune disease | 13 (2.6%) | 1 (0.5%) | 8 (4.0%) | 4 (3.8%) |
| Hypothyroidism | 12 (2.4%) | 5 (2.5%) | 2 (1.0%) | 5 (4.8%) |
| Hyperthyroidism | 8 (1.6%) | 6 (3.0%) | 2 (1.0%) | 0 (0.0%) |
| Addison's disease | 2 (0.4%) | 2 (1.0%) | 0 (0.0%) | 0 (0.0%) |
| None of these | 187 (37.0%) | 89 (44.5%) | 69 (34.5%) | 29 (27.6%) |

MASH, metabolic dysfunction-associated steatohepatitis; MASLD, metabolic dysfunction-associated steatotic liver disease; NASH, non-alcoholic steatohepatitis; NAFLD, non-alcoholic fatty liver disease; SD, standard deviation.

The mean (SD) time since liver disease diagnosis was 3.9 (5.9) years, with the shortest time in India (2.0 [1.8] years) and the highest in China (6.2 [8.3] years) (**Table 1** and **Supplementary Table 3**). The most common type of liver disease was NAFLD/NASH (44.4%), followed by hepatitis B (22.6%), alcohol-related liver disease (15.2%) and MASLD/MASH (12.5%). Note that respondents could select both NAFLD/NASH and MASLD/MASH. There were 273 respondents (54.1%) who selected either NAFLD/NASH or MASLD/MASH or both. Common comorbidities included obesity (26.1%), hyperlipidemia (19.0%), hypertension (18.0%) and anxiety (16.2%). All patients (100%) reported fatigue as per the entrance criteria for the study, while 30.7%, 23.8%, 23.6% and 22.4%, respectively, reported abdominal pain, dark-colored urine, nausea or vomiting and swelling in the arms or legs (**Figure 1**). Many participants considered their fatigue to be moderate (51.3%) or serious (26.9%) in severity, with 33.5% experiencing fatigue every day or almost every day and 48.3% experiencing fatigue 2 to 5 days per week. Two-thirds of participants (63.4%) had discussed their fatigue with a doctor, and none (0%) were receiving treatment for their fatigue.

The most common overall treatments received by participants were antioxidants (18.4%), statins (14.9%) and antiviral drugs (14.1%) (**Supplementary Table 4**). Among participants currently receiving CLD treatment, 48.8% (n=138) were satisfied with their treatment, with 36.4% neither satisfied nor dissatisfied. More than half (56.9%) considered their treatment to have acceptable effectiveness, with 32.2% reporting good effectiveness and just 6.7% reporting very good effectiveness.

Responses on the impact of CLD and fatigue on HRQoL are shown in **Table 2** and **Figure 2**. Just under half of participants reported that their CLD had sometimes, often or always affected their self-esteem/confidence (n=228/505 [45.1%]) or caused embarrassment (n=198/505 [39.2%]) in the prior 6 months, with the greatest impact seen in Mexico (**Figure 2**). Additionally, 33.3% (n=168/505) of participants reported sometimes, often or always experiencing judgement/discrimination due to their CLD in the prior 6 months. Again, this was greatest in Mexico. Ability to maintain or acquire new employment was reported to be sometimes/often/always affected by CLD by 38.6% (n=195/505) of participants, with 22.9% of participants in Mexico reporting that their CLD often affected their ability to acquire new employment compared to just 13.5% in India and 6.5% in China. Three-hundred and forty participants (67.3%) also reported that they worry about the future, with the highest proportion in Mexico (90.5%) followed by India (80.0%) and China (42.5%) (**Table 2**). Participants generally felt supported with their CLD-related fatigue; 67.5% (n=341) were comfortable discussing fatigue with their physician and 72.7% (n=367) reported having support from family and friends with fatigue management. Notably, a higher proportion of participants in China (82.0%) reported being comfortable discussing fatigue with their physician compared to Mexico (64.8%) and India (54.5%). Around half of participants (47.3% [n=239/505]) felt their social life was negatively impacted by their fatigue diagnosis, 41.8% (n=211/505) kept their liver disease a secret, and many said that their CLD-related fatigue had caused financial difficulties (n=272/505 [53.9%]), medical debt (n=148/505 [29.3%]), financial reliance on others (n=172/505 [34.1%]), or reliance on government assistance (n=79/505 [15.6%]). The proportions of participants reporting financial difficulties or medical debt due to CLD-related fatigue were highest in Mexico (76.2% and 57.1%) and lowest in China (30.5% and 12.5%), while reliance on government assistance was highest in India (22.0%) followed by Mexico (17.2%) and China (8.5%).

**Figure 2.**
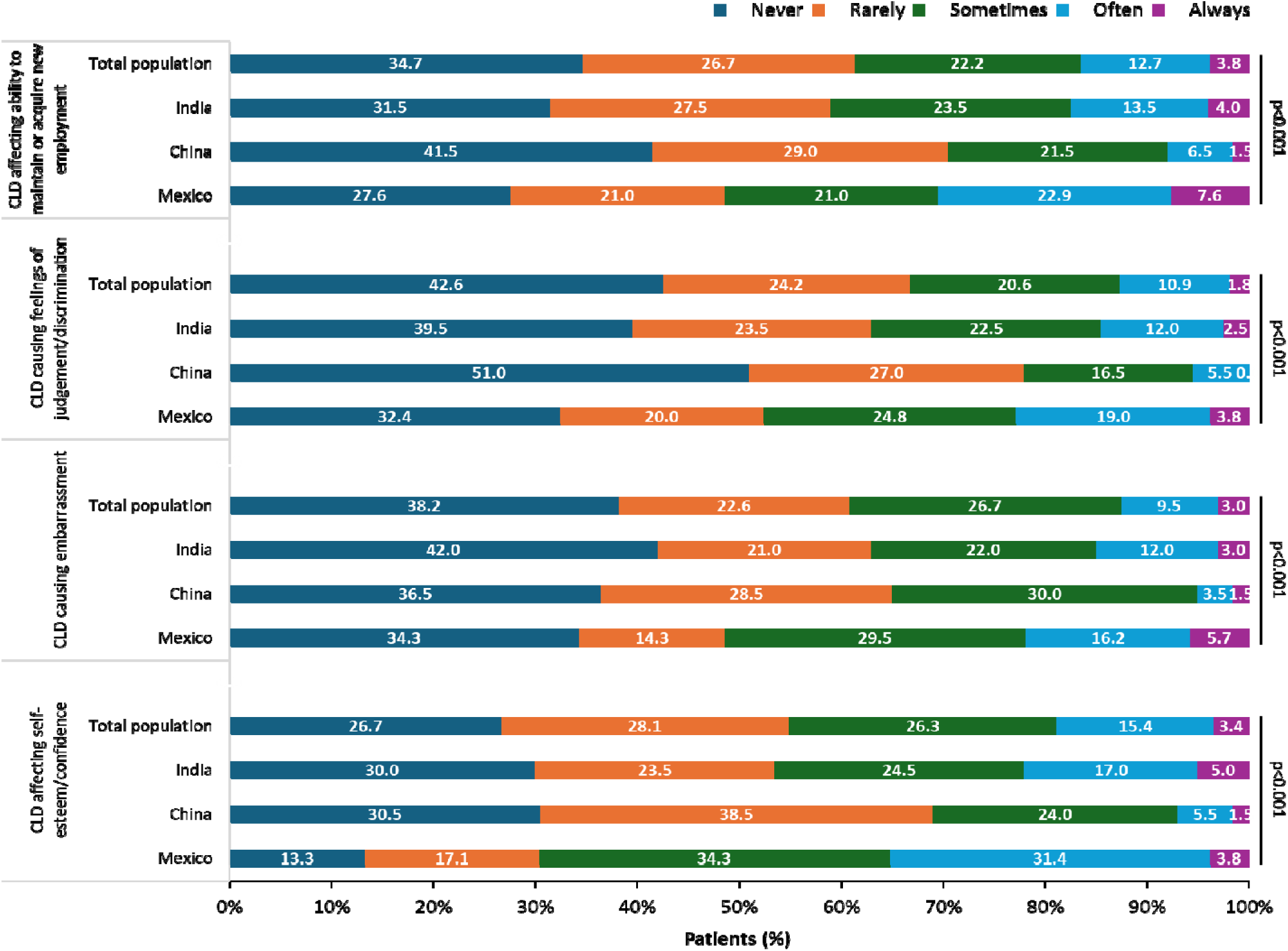
Impact of CLD and fatigue (survey responses) CLD, chronic liver disease.

**Table 2.** Impact of CLD and fatigue (survey responses)

|  | N = 505 | Country |  |  | p-value* |
| --- | --- | --- | --- | --- | --- |
|  |  | India, N = 200 | China, N = 200 | Mexico, N = 105 |  |
| **Worry about future, n (%)** |  |  |  |  | <0.001 |
| Strongly disagree | 17 (3.4%) | 3 (1.5%) | 13 (6.5%) | 1 (1.0%) |  |
| Disagree | 66 (13.1%) | 17 (8.5%) | 47 (23.5%) | 2 (1.9%) |  |
| Uncertain | 81 (16.0%) | 20 (10.0%) | 54 (27.0%) | 7 (6.7%) |  |
| Agree | 215 (42.6%) | 110 (55.0%) | 65 (32.5%) | 40 (38.1%) |  |
| Strongly agree | 125 (24.8%) | 50 (25.0%) | 20 (10.0%) | 55 (52.4%) |  |
| Missing | 1 (0.2%) | 0 (0.0%) | 1 (0.5%) | 0 (0.0%) |  |
| **Comfort discussing liver disease related fatigue with physician, n (%)** |  |  |  |  | <0.001 |
| Strongly disagree | 20 (4.0%) | 15 (7.5%) | 3 (1.5%) | 2 (1.9%) |  |
| Disagree | 58 (11.5%) | 41 (20.5%) | 6 (3.0%) | 11 (10.5%) |  |
| Uncertain | 86 (17.0%) | 35 (17.5%) | 27 (13.5%) | 24 (22.9%) |  |
| Agree | 257 (50.9%) | 89 (44.5%) | 123 (61.5%) | 45 (42.9%) |  |
| Strongly agree | 84 (16.6%) | 20 (10.0%) | 41 (20.5%) | 23 (21.9%) |  |
| **Support from family and friends with fatigue management, n (%)** |  |  |  |  | <0.001 |
| Strongly disagree | 18 (3.6%) | 7 (3.5%) | 1 (0.5%) | 10 (9.5%) |  |
| Disagree | 47 (9.3%) | 28 (14.0%) | 10 (5.0%) | 9 (8.6%) |  |
| Uncertain | 73 (14.5%) | 14 (7.0%) | 40 (20.0%) | 19 (18.1%) |  |
| Agree | 255 (50.5%) | 91 (45.5%) | 123 (61.5%) | 41 (39.0%) |  |
| Strongly agree | 112 (22.2%) | 60 (30.0%) | 26 (13.0%) | 26 (24.8%) |  |
| **Social life negatively impacted due to fatigue diagnosis, n (%)** |  |  |  |  | <0.001 |
| Strongly disagree | 33 (6.5%) | 15 (7.5%) | 16 (8.0%) | 2 (1.9%) |  |
| Disagree | 99 (19.6%) | 41 (20.5%) | 52 (26.0%) | 6 (5.7%) |  |
| Uncertain | 134 (26.5%) | 45 (22.5%) | 64 (32.0%) | 25 (23.8%) |  |
| Agree | 184 (36.4%) | 78 (39.0%) | 59 (29.5%) | 47 (44.8%) |  |
| Strongly agree | 55 (10.9%) | 21 (10.5%) | 9 (4.5%) | 25 (23.8%) |  |
| **Keeping liver disease status a secret, n (%)** |  |  |  |  | 0.002 |
| Strongly disagree | 59 (11.7%) | 20 (10.0%) | 18 (9.0%) | 21 (20.0%) |  |
| Disagree | 126 (25.0%) | 59 (29.5%) | 56 (28.0%) | 11 (10.5%) |  |
| Uncertain | 109 (21.6%) | 39 (19.5%) | 46 (23.0%) | 24 (22.9%) |  |
| Agree | 160 (31.7%) | 68 (34.0%) | 57 (28.5%) | 35 (33.3%) |  |
| Strongly agree | 51 (10.1%) | 14 (7.0%) | 23 (11.5%) | 14 (13.3%) |  |
| **Liver disease related fatigue caused financial difficulties, n (%)** |  |  |  |  | <0.001 |
| Strongly disagree | 69 (13.7%) | 13 (6.5%) | 50 (25.0%) | 6 (5.7%) |  |
| Disagree | 164 (32.5%) | 56 (28.0%) | 89 (44.5%) | 19 (18.1%) |  |
| Agree | 195 (38.6%) | 84 (42.0%) | 59 (29.5%) | 52 (49.5%) |  |
| Strongly agree | 77 (15.2%) | 47 (23.5%) | 2 (1.0%) | 28 (26.7%) |  |
| **Liver disease related fatigue caused medical debt, n (%)** |  |  |  |  | <0.001 |
| Strongly disagree | 131 (25.9%) | 52 (26.0%) | 69 (34.5%) | 10 (9.5%) |  |
| Disagree | 226 (44.8%) | 85 (42.5%) | 106 (53.0%) | 35 (33.3%) |  |
| Agree | 112 (22.2%) | 50 (25.0%) | 22 (11.0%) | 40 (38.1%) |  |
| Strongly agree | 36 (7.1%) | 13 (6.5%) | 3 (1.5%) | 20 (19.0%) |  |
| **Financially reliant on others due to liver disease related fatigue, n (%)** |  |  |  |  | <0.001 |
| Strongly disagree | 151 (29.9%) | 61 (30.5%) | 64 (32.0%) | 26 (24.8%) |  |
| Disagree | 182 (36.0%) | 65 (32.5%) | 89 (44.5%) | 28 (26.7%) |  |
| Agree | 135 (26.7%) | 58 (29.0%) | 44 (22.0%) | 33 (31.4%) |  |
| Strongly agree | 37 (7.3%) | 16 (8.0%) | 3 (1.5%) | 18 (17.1%) |  |
| **Reliance on government assistance due to liver disease related fatigue, n (%)** |  |  |  |  | <0.001 |
| Strongly disagree | 219 (43.4%) | 66 (33.0%) | 101 (50.5%) | 52 (49.5%) |  |
| Disagree | 207 (41.0%) | 90 (45.0%) | 82 (41.0%) | 35 (33.3%) |  |
| Agree | 62 (12.3%) | 30 (15.0%) | 17 (8.5%) | 15 (14.3%) |  |
| Strongly agree | 17 (3.4%) | 14 (7.0%) | 0 (0.0%) | 3 (2.9%) |  |

\*Pearson's Chi-squared test; Chi-Squared Test with Monte Carlo Simulation. CLD, chronic
liver
disease.

Responses relating to medical consultations/hospital visits are shown in **Table 3** and **Figure 3**. In the past 12 months, 75.6% of participants had visited a hepatologist (lowest in India [56.4%] and highest in China [89.1%]), 72.0% had visited a general practitioner (lowest in China [48.9%] and highest in Mexico [96.9%]), 57.0% had visited a gastroenterologist (lowest in China [42.4%] and highest in Mexico [70.4%]), and 50.3% had visited an internal medicine specialist (lowest in India [36.4%] and highest in Mexico [71.4%]) (**Table 3**). There was a mean (SD) of 0.8 (1.6) emergency consultations due to CLD in the 12 months prior to the interview, with a mean (SD) of 2.4 (6.0) days of CLD-related hospitalization (**Figure 3**). The mean number of emergency consultations due to CLD was highest in Mexico (1.5 [2.0]) while the mean number of days hospitalized due to CLD was highest in China (2.7 [6.7]) (**Figure 3**).

**Table 3.** Medical consultations/hospital visits (survey responses)

|  | N = 505 | Country |  |  | p-value* |
| --- | --- | --- | --- | --- | --- |
|  |  | India, N = 200 | China, N = 200 | Mexico, N = 105 |  |
| Physician visits in the past 12 months, n (%) |  |  |  |  |  |
| Hepatologist | 338 (75.6%) | 93 (56.4%) | 164 (89.1%) | 81 (82.7%) | <0.001 |
| General Practitioner | 322 (72.0%) | 137 (83.0%) | 90 (48.9%) | 95 (96.9%) | <0.001 |
| Gastroenterologist | 255 (57.0%) | 108 (65.5%) | 78 (42.4%) | 69 (70.4%) | <0.001 |
| Internist/Internal Medicine Specialist | 225 (50.3%) | 60 (36.4%) | 95 (51.6%) | 70 (71.4%) | <0.001 |
| Other | 25 (44.6%) | 3 (13.6%) | 18 (75.0%) | 4 (40.0%) | <0.001 |
| None | 58 (11.5%) | 35 (17.5%) | 16 (8.0%) | 7 (6.7%) | 0.003 |

\*Pearson's Chi-squared test; Fisher's exact test; Kruskal-Wallis rank sum test.
CLD,
chronic
liver
disease.

**Figure 3.**
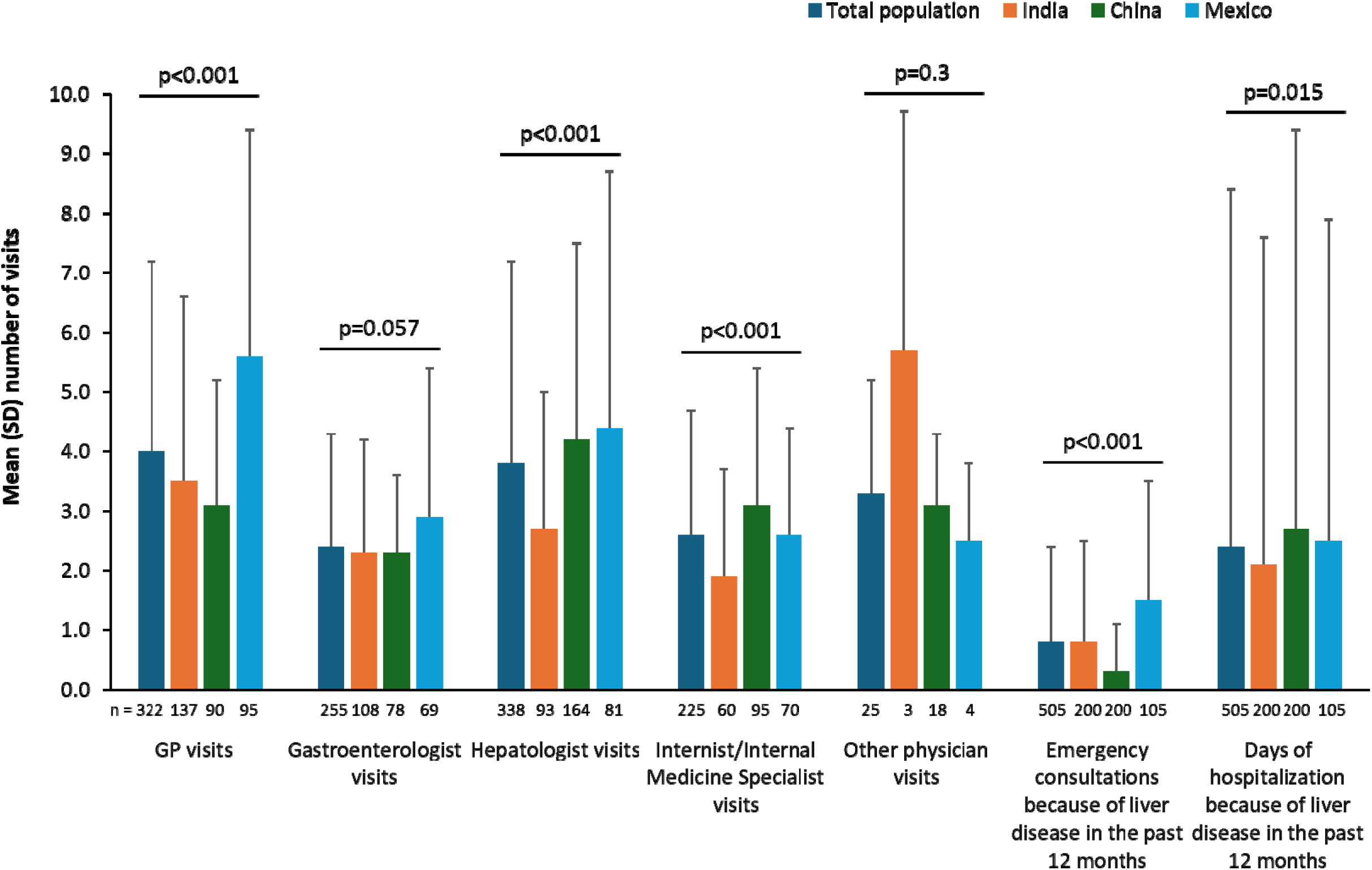
Physician visits in the past 12 months (in patients having at least one visit to the respective physician) GP, general practitioner; SD, standard deviation.

### Patient-Reported Outcomes Measurement Information System-29+2

PROMIS scores are shown in **Table 4**. The highest mean (SD) scores in the total population were for anxiety (59.4 [8.6]), pain interference (57.2 [8.6]), depression (55.9 [9.3]), fatigue (55.5 [9.0]) and sleep disturbance (54.4 [6.5]), all of which were highest in Mexico.

**Table 4.**
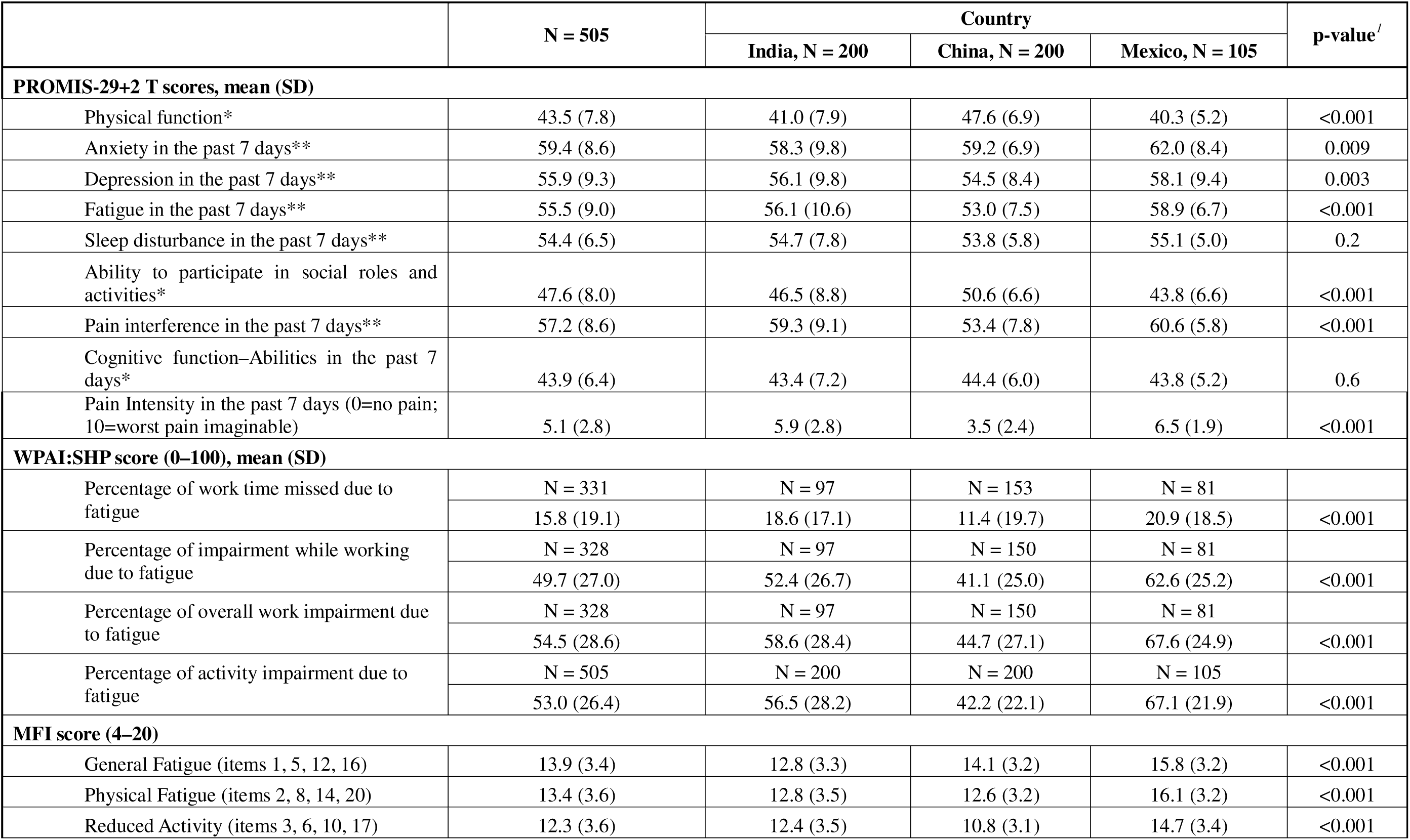
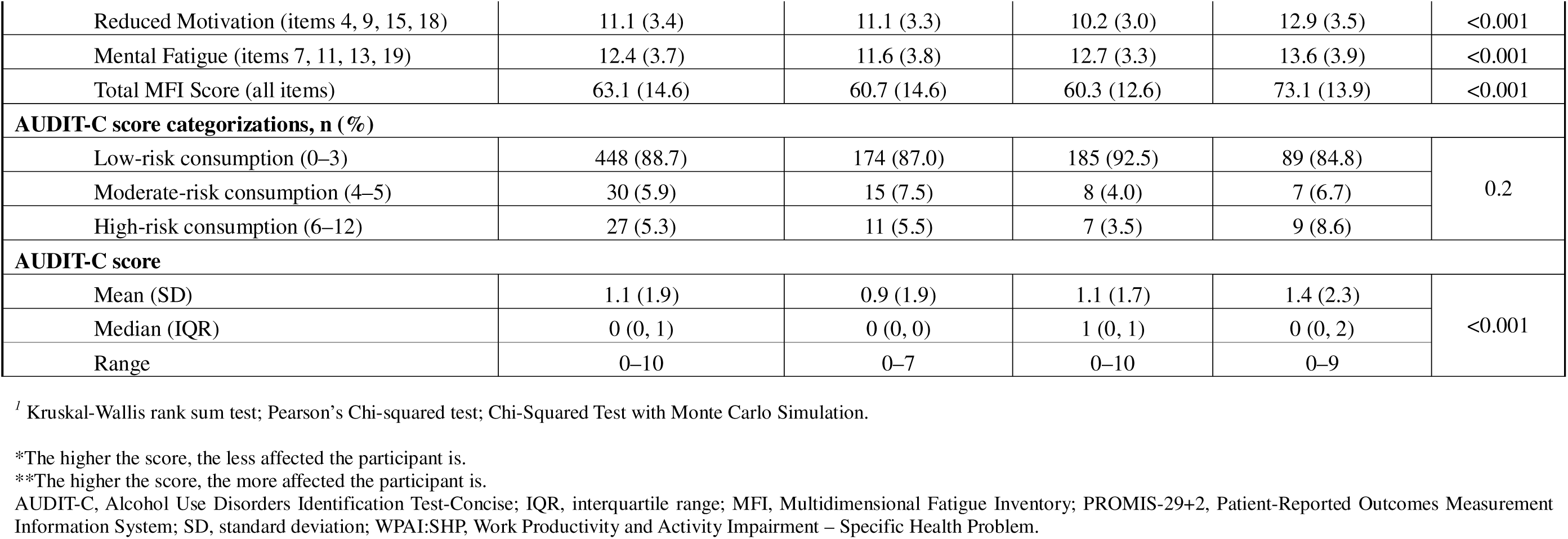
Patient-reported outcome measures.

|  | N = 505 | Country |  |  | p-value<sup>1</sup> |
| --- | --- | --- | --- | --- | --- |
|  |  | India, N = 200 | China, N = 200 | Mexico, N = 105 |  |
| PROMIS-29+2 T scores, mean (SD) |  |  |  |  |  |
| Physical function* | 43.5 (7.8) | 41.0 (7.9) | 47.6 (6.9) | 40.3 (5.2) | <0.001 |
| Anxiety in the past 7 days** | 59.4 (8.6) | 58.3 (9.8) | 59.2 (6.9) | 62.0 (8.4) | 0.009 |
| Depression in the past 7 days** | 55.9 (9.3) | 56.1 (9.8) | 54.5 (8.4) | 58.1 (9.4) | 0.003 |
| Fatigue in the past 7 days** | 55.5 (9.0) | 56.1 (10.6) | 53.0 (7.5) | 58.9 (6.7) | <0.001 |
| Sleep disturbance in the past 7 days** | 54.4 (6.5) | 54.7 (7.8) | 53.8 (5.8) | 55.1 (5.0) | 0.2 |
| Ability to participate in social roles and activities* | 47.6 (8.0) | 46.5 (8.8) | 50.6 (6.6) | 43.8 (6.6) | <0.001 |
| Pain interference in the past 7 days** | 57.2 (8.6) | 59.3 (9.1) | 53.4 (7.8) | 60.6 (5.8) | <0.001 |
| Cognitive function–Abilities in the past 7 days* | 43.9 (6.4) | 43.4 (7.2) | 44.4 (6.0) | 43.8 (5.2) | 0.6 |
| Pain Intensity in the past 7 days (0=no pain; 10=worst pain imaginable) | 5.1 (2.8) | 5.9 (2.8) | 3.5 (2.4) | 6.5 (1.9) | <0.001 |
| WPAI:SHP score (0–100), mean (SD) |  |  |  |  |  |
| Percentage of work time missed due to fatigue | N = 331 | N = 97 | N = 153 | N = 81 |  |
|  | 15.8 (19.1) | 18.6 (17.1) | 11.4 (19.7) | 20.9 (18.5) | <0.001 |
| Percentage of impairment while working due to fatigue | N = 328 | N = 97 | N = 150 | N = 81 |  |
|  | 49.7 (27.0) | 52.4 (26.7) | 41.1 (25.0) | 62.6 (25.2) | <0.001 |
| Percentage of overall work impairment due to fatigue | N = 328 | N = 97 | N = 150 | N = 81 |  |
|  | 54.5 (28.6) | 58.6 (28.4) | 44.7 (27.1) | 67.6 (24.9) | <0.001 |
| Percentage of activity impairment due to fatigue | N = 505 | N = 200 | N = 200 | N = 105 |  |
|  | 53.0 (26.4) | 56.5 (28.2) | 42.2 (22.1) | 67.1 (21.9) | <0.001 |
| MFI score (4–20) |  |  |  |  |  |
| General Fatigue (items 1, 5, 12, 16) | 13.9 (3.4) | 12.8 (3.3) | 14.1 (3.2) | 15.8 (3.2) | <0.001 |
| Physical Fatigue (items 2, 8, 14, 20) | 13.4 (3.6) | 12.8 (3.5) | 12.6 (3.2) | 16.1 (3.2) | <0.001 |
| Reduced Activity (items 3, 6, 10, 17) | 12.3 (3.6) | 12.4 (3.5) | 10.8 (3.1) | 14.7 (3.4) | <0.001 |
| Reduced Motivation (items 4, 9, 15, 18) | 11.1 (3.4) | 11.1 (3.3) | 10.2 (3.0) | 12.9 (3.5) | <0.001 |
| Mental Fatigue (items 7, 11, 13, 19) | 12.4 (3.7) | 11.6 (3.8) | 12.7 (3.3) | 13.6 (3.9) | <0.001 |
| Total MFI Score (all items) | 63.1 (14.6) | 60.7 (14.6) | 60.3 (12.6) | 73.1 (13.9) | <0.001 |
| AUDIT-C score categorizations, n (%) |  |  |  |  |  |
| Low-risk consumption (0–3) | 448 (88.7) | 174 (87.0) | 185 (92.5) | 89 (84.8) | 0.2 |
| Moderate-risk consumption (4–5) | 30 (5.9) | 15 (7.5) | 8 (4.0) | 7 (6.7) |  |
| High-risk consumption (6–12) | 27 (5.3) | 11 (5.5) | 7 (3.5) | 9 (8.6) |  |
| AUDIT-C score |  |  |  |  |  |
| Mean (SD) | 1.1 (1.9) | 0.9 (1.9) | 1.1 (1.7) | 1.4 (2.3) | <0.001 |
| Median (IQR) | 0 (0, 1) | 0 (0, 0) | 1 (0, 1) | 0 (0, 2) |  |
| Range | 0–10 | 0–7 | 0–10 | 0–9 |  |

<sup>1</sup> Kruskal-Wallis rank sum test; Pearson's Chi-squared test; Chi-Squared Test with Monte Carlo Simulation.
\*The higher the score, the less affected the participant is.
\*\*The higher the score, the more affected the participant is.
AUDIT-C, Alcohol Use Disorders Identification Test-Concise; IQR, interquartile range; MFI, Multidimensional Fatigue Inventory; PROMIS-29+2, Patient-Reported Outcomes Measurement Information System; SD, standard deviation; WPAI:SHP, Work Productivity and Activity Impairment – Specific Health Problem.

### Work Productivity and Activity Impairment – Specific Health Problem

The mean (SD) WPAI:SHP score, assessing activity impairment due to health problems, was 53.0 (26.4). Scores were highest (indicating greater impairment) in Mexico (67.1 [21.9]), followed by India (56.5 [28.2]) and China (42.2 [22.1]) (**Table 4**). For those who reported hours actually worked, the mean (SD) WPAI:SHP scores for percentage impairment while working was 49.7 (27.0), 54.5 (28.6) for percentage of overall work impairment and 15.8 (19.1) for percentage of work time missed due to fatigue.

### Multidimensional Fatigue Inventory

The mean (SD) total MFI score was 63.1 (14.6) for the overall population, with the highest score seen in Mexico (73.1 [13.9]) (**Table 4**). Mean domain scores for general fatigue, physical fatigue, reduced activity, reduced motivation, and mental fatigue were also highest in Mexico (**Table 4**).

### Alcohol Use Disorders Identification Test-Concise

AUDIT-C scores were generally in the low-risk consumption category (88.7% of the total population) and this was similar across India (87.0%), China (92.5%) and Mexico (84.8%; **Table 4**).

### Subgroup analyses

In general, the proportion of patients with fatigue, PROMIS-29+2 scores, WPAI:SHP scores and MFI scores were similar across patient subgroups analyzed (**Supplementary Tables 7-10**). However, mental fatigue was higher among patients with longer CLD duration and among patients who were currently or had previously been treated for CLD compared with those who had never been treated (**Supplementary Table 7**). Additionally, PROMIS-29+2 scores tended to be higher in females than in males and in patients with higher versus lower fatigue severity (**Supplementary Table 8**). WPAI:SHP scores were consistently higher among patients with alcohol-related liver disease versus others and in patients with higher versus lower fatigue severity (**Supplementary Table 9**). Finally, MFI scores were consistently higher in females than in males and in patients with alcohol-related liver disease versus others (**Supplementary Table 10**).

## Discussion

This multinational, cross-sectional study provides important data demonstrating the substantial patient-reported impact of CLD-related fatigue on daily activities and HRQoL across China, Mexico and India. These countries were selected due to the high prevalence of CLD reported, with particularly high rates of mortality and disability-adjusted life years due to metabolic dysfunction-associated steatotic liver disease compared to other countries globally.^16^ To our knowledge, this study represents the first multinational study reporting patient-reported outcomes for patients with CLD focusing on fatigue as a central symptom. By study design, participants receiving pharmacologic treatments specifically prescribed for fatigue were excluded; thus, none of the included individuals were on fatigue targeted therapies. Most participants reported that their fatigue was moderate or serious in severity, with one-third of participants experiencing fatigue every day or almost every day and around half reporting that their fatigue negatively impacted their social life. In addition, over half of participants reported that their CLD-related fatigue had caused financial difficulties and more than one-third said their ability to maintain or acquire new employment was impacted. Medical consultations and hospital visits were also frequent. Use of validated PRO tools demonstrated severe fatigue (MFI score ≥63) in the population as well as substantial levels of work and activity impairment (WPAI:SHP scores ≥67) and high levels of anxiety, pain interference, depression and sleep interference (PROMIS scores ≥54). Together, these data highlight the need for effective identification and management of CLD-related fatigue to reduce the sizeable clinical and societal burden and to improve patients’ day-to-day lives.

CLD-related fatigue is common among patients with CLD, with prevalence estimates ranging between 50% and 85%.^5^ As such, it is vital that fatigue and its impact are better understood, particularly from the patient’s point of view. Our data build on data from previous studies that have demonstrated the patient-reported impact of CLD-related fatigue. For example, another survey study investigating if the type and severity of CLD had different effects on HRQoL found that chronic fatigue was prevalent in patients with cholestatic liver disease and had an important impact on their HRQoL.^17^ Indeed, this study found HRQoL of patients with CLD to be similar to patients with chronic obstructive pulmonary disease or congestive heart failure.^17^ Typically, fatigue does not correlate with CLD severity, with patients who have earlier stage CLD also experiencing a degree of fatigue that causes negative HRQoL impacts.^7^ Fatigue was present in all study participants, as per the inclusion criteria, and notably, the study cohort included a range of patients with different CLD types and differing disease durations. Approximately 15% of patients included in the study were prescribed statins, most probably due to dyslipidemia, which is an etiologic co-factor in MASLD. Fatigue is a recognized side effect of statins, often characterized as muscle pain, weakness or reduced energy and referred to as statin-associated muscle symptoms.^18^ Although these medications were not prescribed for fatigue, their use may contribute to fatigue symptoms and therefore represents a potential confounder. As the medication lists were self-reported by participants and not verified through medical records, we could not determine whether lipid-lowering therapy contributed to fatigue severity in this cohort.

Therefore, fatigue assessment may be warranted in routine care for all patients with CLD to support timely identification and implementation of evidence-based supportive strategies where necessary.

Interestingly, there were some country-level differences in fatigue perception and support in the current study that likely relate to differences in sociodemographic and clinical characteristics across countries. For example, full-time employment was much more common in China than in Mexico and India, as was being educated to a college/university level. Higher rates of full-time employment in the Chinese sample may also have reduced concerns about job security relative to participants in India and Mexico. Education status is likely to impact disease understanding and management, and may explain the higher levels of worry about the future reported among participants in Mexico and India. Moreover, compared to their counterparts in China, patients in Mexico and India were less likely to be comfortable discussing fatigue with their physician. However, it is likely that the proportion of participants reporting being comfortable discussing fatigue with their physician is artificially high in some areas due to participation bias, as many patients were recruited to the study through physician referral, particularly in China (61.0%) and India (55.5%). The proportion of participants experiencing judgement and discrimination due to their CLD-related fatigue was highest in Mexico, which may explain why more patients in Mexico reported experiencing problems with maintaining or acquiring employment. While there are challenges related to the management of CLD-related fatigue in clinical practice, supportive strategies are known to reduce fatigue burden. Moreover, advances in our understanding of the mechanisms underlying fatigue in CLD have the potential to lead to future specific, targeted therapies.^6,19^ As such, it is important for physicians to actively assess for presence and impact of fatigue in patients with CLD to facilitate effective identification and treatment, particularly given the wide availability of fatigue assessment tools.^6,20^ Notably, among patients receiving treatment for their CLD, most participants (95.8%) considered their current CLD treatment to have either acceptable, good or very good effectiveness, while only around half (56.9%) were satisfied (or very satisfied) with their treatment. This finding suggests that factors other than perceived treatment effectiveness are also important to patients, emphasizing the importance of shared decision-making in CLD to ensure patient views are considered when developing a management strategy.

This study has several strengths including the large participant population, its multinational nature and the use of well-known, validated PRO tools to measure CLD-related fatigue and its impact. Limitations to note include that participants had a self-reported fatigue diagnosis that was not directly validated through clinical data which may introduce selection (misclassification) bias. This study does not take into account the differences in access to healthcare in different countries or variations in practice. Sampling and recruitment through physician referral, patient associations, and pre existing research panels may have introduced selection and participation biases, potentially over representing individuals with higher healthcare engagement, digital literacy, or closer links to medical providers. This is particularly relevant given the high proportion of participants referred by physicians in China and India, which could inflate country level estimates of comfort discussing fatigue with clinicians and should temper interpretation of cross country comparisons. Additionally, the study had a cross-sectional design which may make it susceptible to responder and recall bias. There are some limitations regarding PRO instruments: for the MFI 20, there are no universally standardized severity thresholds. We report continuous scores as primary and used a ≥60 total score threshold exploratorily to describe “high burden fatigue,” consistent with prior psychometric work proposing cut point–based classification; nonetheless, any thresholding should be viewed as heuristic.^4^ By design, all participants were experiencing fatigue at enrolment, so the study cannot quantify how much of the observed HRQoL, work and psychosocial burden is specifically attributable to fatigue versus to CLD itself. Finally, the impacts attributed to fatigue (e.g., social, emotional, financial, work-related) were self-reported and not corroborated by clinical assessments or objective records, and therefore may be subject to reporting or attribution bias. Despite these limitations, the study provides important insights into the day-to-day patient-reported experiences and impact of living with CLD-related fatigue that are crucial for clinicians to be aware of when treating their patients. Together with previous studies demonstrating patient-reported CLD impacts, this study highlights a global need for improved understanding, awareness and management of CLD-related fatigue.

## Conclusions

In this multinational cohort of adults with compensated chronic liver disease, fatigue was highly prevalent, persistent, and associated with substantial impairment in HRQoL, daily functioning, and psychological well being. These findings demonstrate a clear global unmet need for systematic assessment and targeted management of fatigue in routine CLD care, alongside supportive strategies that address both the physical and psychological dimensions of this burdensome symptom.

## Supporting information

CLD GAP Study Manuscript supplement

## Abbreviations

aGAP: Abbott Global Assessment of Patients unmet needs
ANOVA: analysis of variance
AUDIT-C: Alcohol Use Disorders Identification Test-Concise
CLD: chronic liver disease
HRQoL: health-related quality of life
IQR: interquartile range
MASH: metabolic dysfunction-associated steatohepatitis
MASLD: metabolic dysfunction-associated steatotic liver disease
MFI: Multidimensional Fatigue Inventory
NASH: non-alcoholic steatohepatitis
NAFLD: non-alcoholic fatty liver disease
PRO: patient-reported outcome
PROM: patient-reported outcome measures
PROMIS-29+2: Patient-Reported Outcomes Measurement Information System
SD: standard deviation
WPAI:SHP: Work Productivity and Activity Impairment – Specific Health Problem.

## Data Availability

All relevant data are within the paper and its Supporting Information files.

## Acknowledgements

Medical writing and editorial assistance were provided by Martin Guppy PhD of Metamols Ltd, funded by Abbott Established Pharmaceuticals, in accordance with Good Publication Practice (GPP 2022) guidelines.

## Author contributions

GC, GA-U, NN, MMC, XG, RGD, AS, and MGS, contributed equally to the conception, design and writing of the manuscript. All critically revised the manuscript, agreed to be fully accountable for ensuring the integrity and accuracy of the work, and read and approved the final manuscript.

