## Supplementary material for "Burden of fatigue in compensated chronic liver disease: findings from the multinational a:GAP Study": CLD GAP Study Manuscript supplement

**a:GAP Study** **Supplementary Tables**

**Supplementary Table 1**. Search Algorithm

Database details:

Journals@Ovid Full Text <October 23, 2023>

Your Journals@Ovid

EB Health - KSR Evidence <2015 to 2023 Week 42>

EB Health - ECRI Guidelines Trust <2018 to 2023 Week 42>

EB Health - Family Physicians Inquiries Network : Evidence-Based Practice <2013 to October 2023>

EB Health - Lippincott Advisor Clinical Guideline Summaries <2018 to October 2023>

Ovid MEDLINE(R) ALL <1946 to October 23, 2023>

| **#** | **Query** | **Results (24 October 2023**) |
| --- | --- | --- |
| 1 | Chronic liver disease or CLD or cirrhosis or nonalcoholic fatty liver disease or NALFD or nonalcoholic steatohepatitis or NASH or alcoholic liver disease or hepatitis B virus liver disease or hepatitis C virus liver disease or autoimmune hepatitis or primary sclerosing cholangitis or primary biliary cholangitis or Wilson's disease or Alpha-1-antitrypsin deficiency or haemochromatosis or hepatic steatosis or cystic fibrosis-related liver disease or nodular regenerative hyperplasia | 233,051 |
| 2 | Symptoms or fatigue | 2,022.379 |
| 3 | 1 and 2 | 11,959 |
| 4 | Symptom burden or fatigue burden or symptom impact or fatigue impact. | 9 102 |
| 5 | Cost burden or economic burden or financial burden or cost impact or economic impact or financial impact | 52 089 |
| 6 | Humanistic burden or health related quality of life or HRQoL or quality of life or QOL or functionality or utility or productivity | 952,196 |
| 7 | Treatment or management or care | 8,248,119 |
| 8 | Satisfaction or goals or success or preferences or priorities or communication or decision making or shared decision making | 1,454, 133 |
| 9 | 7 and 8 | 656,273 |
| 10 | 3 and 4 | 149 |
| 11 | 3 and 5 | 25 |
| 12 | 3 and 6 | 1,041 |
| 13 | 3 and 9 | 293 |
| 14 | Quantitative or qualitative or mixed methods or surveys or questionnaires or interviews or focus group or registry or cohorts. | 4,667,432 |
| 15 | 10 and 14 | 88 |
| 16 | 11 and 14 | 8 |
| 17 | 12 and 14 | 511 |
| 18 | 14 and 14 | 76 |
| 19 | Remove duplicates from 15 | 65 |
| 20 | Remove duplicates from 16 | 6 |
| 21 | Remove duplicates from 17 | 367 |
| 22 | Remove duplicates from 18 | 56 |

CLD, chronic liver disease; HRQoL, health-related quality of life; NASH, non-alcoholic steatohepatitis; NAFLD, non-alcoholic fatty liver disease; QoL, quality of life.

**Supplementary Table 2.** Sociodemographic information

| **Base: All participants** | **N = 505** | **Country** | | | **p-value***^1^* |
| --- | --- | --- | --- | --- | --- |
|  |  | **India, N = 200** | **China, N = 200** | **Mexico, N = 105** |  |
| **S1: Age** | | | | | 0.002 |
| N | 505 | 200 | 200 | 105 |  |
| Mean (SD) | 41.3 (12.6) | 39.4 (12.9) | 43.5 (11.9) | 40.5 (12.7) |  |
| Median (IQR) | 40 (32, 49) | 38 (30, 47) | 42 (35, 50) | 40 (29, 50) |  |
| Range | 18–82 | 18–82 | 20–72 | 19–68 |  |
| **S3: Sex, n (%)** | | | | | 0.067 |
| Male | 268 (53.1%) | 110 (55.0%) | 113 (56.5%) | 45 (42.9%) |  |
| Female | 236 (46.7%) | 90 (45.0%) | 86 (43.0%) | 60 (57.1%) |  |
| Prefer not to answer | 1 (0.2%) | 0 (0.0%) | 1 (0.5%) | 0 (0.0%) |  |
| **E4: Marital status, n (%)** | | | | | <0.001 |
| Married | 355 (70.3%) | 153 (76.5%) | 165 (82.5%) | 37 (35.2%) |  |
| Single, never married | 101 (20.0%) | 43 (21.5%) | 23 (11.5%) | 35 (33.3%) |  |
| Divorced | 12 (2.4%) | 0 (0.0%) | 3 (1.5%) | 9 (8.6%) |  |
| Separated | 8 (1.6%) | 0 (0.0%) | 0 (0.0%) | 8 (7.6%) |  |
| Widowed | 7 (1.4%) | 3 (1.5%) | 1 (0.5%) | 3 (2.9%) |  |
| Living with partner | 19 (3.8%) | 0 (0.0%) | 6 (3.0%) | 13 (12.4%) |  |
| Prefer not to answer | 3 (0.6%) | 1 (0.5%) | 2 (1.0%) | 0 (0.0%) |  |
| **E5: Household composition, n (%)** | | | | | 0.065 |
| Living in a regular house/apartment | 499 (98.8%) | 196 (98.0%) | 199 (99.5%) | 104 (99.0%) |  |
| Living in an assisted living facility | 4 (0.8%) | 4 (2.0%) | 0 (0.0%) | 0 (0.0%) |  |
| Living in a nursing home or a care facility | 0 (0.0%) | 0 (0.0%) | 0 (0.0%) | 0 (0.0%) |  |
| Other | 2 (0.4%) | 0 (0.0%) | 1 (0.5%) | 1 (1.0%) |  |
| **E6: Current living situation (multiple responses possible), n (%)** | | | | | |
| Living with a partner/spouse | 339 (67.7%) | 130 (66.3%) | 161 (80.5%) | 48 (45.7%) | <0.001 |
| Living with children | 166 (33.1%) | 84 (42.9%) | 62 (31.0%) | 20 (19.0%) | <0.001 |
| Living with parents or friends | 86 (17.2%) | 45 (23.0%) | 19 (9.5%) | 22 (21.0%) | <0.001 |
| Living alone | 37 (7.4%) | 1 (0.5%) | 14 (7.0%) | 22 (21.0%) | <0.001 |
| Other | 9 (1.8%) | 2 (1.0%) | 2 (1.0%) | 5 (4.8%) | 0.044 |
| **E7: Number of adults in the household (on those who are living in a regular house/apartment or in other household composition)** | | | | | <0.001 |
| N | 499 | 195 | 199 | 105 |  |
| Mean (SD) | 2.9 (1.4) | 3.5 (1.5) | 2.6 (1.1) | 2.2 (1.2) |  |
| Median (IQR) | 2 (2, 4) | 3 (2, 4) | 2 (2, 3) | 2 (1, 3) |  |
| Range | 1–9 | 1–9 | 1–8 | 1–9 |  |
| Missing | 2 (0.4%) | 1 (0.5%) | 1 (0.5%) | 0 (0.0%) |  |
| **E8: Number of children (under 18 years old) in the household (on those who mentioned living with children)** | | | | | 0.2 |
| N | 166 | 84 | 62 | 20 |  |
| Mean (SD) | 1.2 (0.8) | 1.2 (0.9) | 1.0 (0.7) | 1.4 (0.8) |  |
| Median (IQR) | 1 (1, 2) | 1 (1, 2) | 1 (1, 1) | 1 (1, 2) |  |
| Range | 0–4 | 0–4 | 0–3 | 0–3 |  |
| **E9: Education, n (%)** | | | | | <0.001 |
| Less than high school | 35 (6.9%) | 14 (7.0%) | 13 (6.5%) | 8 (7.6%) |  |
| High school | 121 (24.0%) | 44 (22.0%) | 45 (22.5%) | 32 (30.5%) |  |
| College/university | 281 (55.6%) | 87 (43.5%) | 135 (67.5%) | 59 (56.2%) |  |
| Postgraduate | 67 (13.3%) | 55 (27.5%) | 7 (3.5%) | 5 (4.8%) |  |
| Decline to answer | 1 (0.2%) | 0 (0.0%) | 0 (0.0%) | 1 (1.0%) |  |
| **E10: Current employment status, n (%)** | | | | | <0.001 |
| Employed (full time) | 242 (47.9%) | 69 (34.5%) | 142 (71.0%) | 31 (29.5%) |  |
| Employed (part time) | 54 (10.7%) | 15 (7.5%) | 4 (2.0%) | 35 (33.3%) |  |
| Self-employed | 44 (8.7%) | 16 (8.0%) | 8 (4.0%) | 20 (19.0%) |  |
| Homemaker | 70 (13.9%) | 63 (31.5%) | 4 (2.0%) | 3 (2.9%) |  |
| Retired | 31 (6.1%) | 3 (1.5%) | 27 (13.5%) | 1 (1.0%) |  |
| Student | 17 (3.4%) | 11 (5.5%) | 1 (0.5%) | 5 (4.8%) |  |
| Not employed | 22 (4.4%) | 10 (5.0%) | 7 (3.5%) | 5 (4.8%) |  |
| Long-term sick leave or long-term disability | 2 (0.4%) | 1 (0.5%) | 1 (0.5%) | 0 (0.0%) |  |
| Other | 2 (0.4%) | 0 (0.0%) | 1 (0.5%) | 1 (1.0%) |  |
| Erroneous data* | 21 (4.2%) | 12 (6.0%) | 5 (2.5%) | 4 (3.8%) |  |
| **E11: Employment status before the diagnosis of liver disease, n (%)** | | | | | <0.001 |
| Employed (full time) | 304 (60.2%) | 84 (42.0%) | 157 (78.5%) | 63 (60.0%) |  |
| Employed (part time) | 34 (6.7%) | 14 (7.0%) | 6 (3.0%) | 14 (13.3%) |  |
| Self-employed | 41 (8.1%) | 17 (8.5%) | 4 (2.0%) | 20 (19.0%) |  |
| Homemaker | 62 (12.3%) | 60 (30.0%) | 1 (0.5%) | 1 (1.0%) |  |
| Retired | 17 (3.4%) | 0 (0.0%) | 17 (8.5%) | 0 (0.0%) |  |
| Student | 31 (6.1%) | 15 (7.5%) | 10 (5.0%) | 6 (5.7%) |  |
| Not employed | 12 (2.4%) | 7 (3.5%) | 4 (2.0%) | 1 (1.0%) |  |
| Long-term sick leave or long- term disability | 0 (0.0%) | 0 (0.0%) | 0 (0.0%) | 0 (0.0%) |  |
| Other | 4 (0.8%) | 3 (1.5%) | 1 (0.5%) | 0 (0.0%) |  |
| *^1^*Kruskal-Wallis rank sum test; Fisher’s exact test; Pearson’s Chi-squared test; Chi-Squared Test with Monte Carlo Simulation. | | | | | |
| * Data were removed due to inconsistencies between answers in E10 and in B3.  IQR, interquartile range; SD, standard deviation. | | | | | |

**Supplementary Table 3**. Clinical data

|  | **N = 505** | **Country** | | | **p-value***^1^* |
| --- | --- | --- | --- | --- | --- |
|  |  | **India, N = 200** | **China, N = 200** | **Mexico, N = 105** |  |
| **Diagnosis of liver disease, n (%)** | | | | | |
| Liver disease | 505 (100.0%) | 200 (100.0%) | 200 (100.0%) | 105 (100.0%) | >0.9 |
| Cardiovascular disease | 22 (4.4%) | 10 (5.0%) | 8 (4.0%) | 4 (3.8%) | 0.9 |
| Cancer | 9 (1.8%) | 0 (0.0%) | 6 (3.0%) | 3 (2.9%) | 0.027 |
| CKD | 4 (0.8%) | 1 (0.5%) | 2 (1.0%) | 1 (1.0%) | >0.9 |
| HIV/AIDS | 2 (0.4%) | 0 (0.0%) | 1 (0.5%) | 1 (1.0%) | 0.7 |
| Asthma | 0 (0.0%) | 0 (0.0%) | 0 (0.0%) | 0 (0.0%) | >0.9 |
| Hemophilia | 0 (0.0%) | 0 (0.0%) | 0 (0.0%) | 0 (0.0%) | >0.9 |
| None of these | 0 (0.0%) | 0 (0.0%) | 0 (0.0%) | 0 (0.0%) | >0.9 |
| **Type of liver disease, n (%)** | | | | | |
| NAFLD/NASH | 224 (44.4%) | 159 (79.5%) | 31 (15.5%) | 34 (32.4%) | <0.001 |
| Hepatitis B | 114 (22.6%) | 11 (5.5%) | 89 (44.5%) | 14 (13.3%) | <0.001 |
| Alcohol-related liver disease | 77 (15.2%) | 23 (11.5%) | 22 (11.0%) | 32 (30.5%) | <0.001 |
| MASLD/MASH | 63 (12.5%) | 13 (6.5%) | 31 (15.5%) | 19 (18.1%) | 0.004 |
| Hepatitis C | 30 (5.9%) | 5 (2.5%) | 17 (8.5%) | 8 (7.6%) | 0.029 |
| Cancer | 19 (3.8%) | 0 (0.0%) | 18 (9.0%) | 1 (1.0%) | <0.001 |
| Autoimmune disease | 8 (1.6%) | 1 (0.5%) | 6 (3.0%) | 1 (1.0%) | 0.14 |
| Genetic disease | 2 (0.4%) | 2 (1.0%) | 0 (0.0%) | 0 (0.0%) | 0.4 |
| Other disease | 14 (2.8%) | 5 (2.5%) | 1 (0.5%) | 8 (7.6%) | 0.003 |
| I don't know / I am not sure | 3 (0.6%) | 1 (0.5%) | 2 (1.0%) | 0 (0.0%) | 0.8 |
| **NAFLD/NASH/MASLD/MASH, n (%)** | | | | | <0.001 |
| No | 232 (45.9%) | 38 (19.0%) | 141 (70.5%) | 53 (50.5%) |  |
| Yes | 273 (54.1%) | 162 (81.0%) | 59 (29.5%) | 52 (49.5%) |  |
| NAFLD/NASH and MASLD/MASH | 16 (3.2%) | 10 (5.0%) | 3 (1.5%) | 3 (2.9%) |  |
| NAFLD/NASH only | 208 (41.2%) | 149 (74.5%) | 28 (14.0%) | 31 (29.5%) |  |
| MASLD/MASH only | 47 (9.3%) | 3 (1.5%) | 28 (14.0%) | 16 (15.2%) |  |
| Steatohepatitis provided in text field | 2 (0.4%) | 0 (0.0%) | 0 (0.0%) | 2 (1.9%) |  |
| **E2: Comorbidities (multiple responses possible), n (%)** | | | | | |
| Obesity | 132 (26.1%) | 54 (27.0%) | 41 (20.5%) | 37 (35.2%) | 0.020 |
| Hyperlipidemia | 96 (19.0%) | 12 (6.0%) | 62 (31.0%) | 22 (21.0%) | <0.001 |
| Hypertension | 91 (18.0%) | 28 (14.0%) | 45 (22.5%) | 18 (17.1%) | 0.084 |
| Anxiety | 82 (16.2%) | 24 (12.0%) | 25 (12.5%) | 33 (31.4%) | <0.001 |
| Depression | 56 (11.1%) | 27 (13.5%) | 9 (4.5%) | 20 (19.0%) | <0.001 |
| Diabetes | 43 (8.5%) | 26 (13.0%) | 9 (4.5%) | 8 (7.6%) | 0.009 |
| Anemia | 26 (5.1%) | 12 (6.0%) | 6 (3.0%) | 8 (7.6%) | 0.2 |
| Malnutrition and Vitamin B deficiency | 25 (5.0%) | 8 (4.0%) | 11 (5.5%) | 6 (5.7%) | 0.7 |
| Sleep apnea | 21 (4.2%) | 10 (5.0%) | 3 (1.5%) | 8 (7.6%) | 0.022 |
| Electrolyte imbalance | 19 (3.8%) | 10 (5.0%) | 2 (1.0%) | 7 (6.7%) | 0.014 |
| Restless legs syndrome | 17 (3.4%) | 14 (7.0%) | 0 (0.0%) | 3 (2.9%) | <0.001 |
| Auto-immune disease | 13 (2.6%) | 1 (0.5%) | 8 (4.0%) | 4 (3.8%) | 0.040 |
| Hypothyroidism | 12 (2.4%) | 5 (2.5%) | 2 (1.0%) | 5 (4.8%) | 0.11 |
| Hyperthyroidism | 8 (1.6%) | 6 (3.0%) | 2 (1.0%) | 0 (0.0%) | 0.11 |
| Addison's disease | 2 (0.4%) | 2 (1.0%) | 0 (0.0%) | 0 (0.0%) | 0.4 |
| None of these | 187 (37.0%) | 89 (44.5%) | 69 (34.5%) | 29 (27.6%) | 0.009 |
| **A1: Number of years since initial liver disease diagnosis** | | | | | <0.001 |
| N | 494 | 198 | 193 | 103 |  |
| Mean (SD) | 3.9 (5.9) | 2.0 (1.8) | 6.2 (8.3) | 3.1 (3.4) |  |
| Median (IQR) | 2 (1, 4) | 2 (1, 3) | 3 (1, 7) | 2 (1, 5) |  |
| Range | 0–42 | 0–8 | 0–42 | 0–20 |  |
| I do not recall, n (%) | 11 (2.2%) | 2 (1.0%) | 7 (3.5%) | 2 (1.9%) |  |
| **S6A: CLD symptoms at diagnosis (multiple responses possible), n (%)** | | | | | |
| Fatigue that is not related to an effort and which persists despite sleep and rest | 445 (88.1%) | 191 (95.5%) | 160 (80.0%) | 94 (89.5%) | <0.001 |
| Abdominal pain | 205 (40.6%) | 77 (38.5%) | 60 (30.0%) | 68 (64.8%) | <0.001 |
| Nausea or vomiting | 175 (34.7%) | 77 (38.5%) | 55 (27.5%) | 43 (41.0%) | 0.022 |
| Dark-colored urine | 150 (29.7%) | 50 (25.0%) | 59 (29.5%) | 41 (39.0%) | 0.039 |
| Swelling in the arms or legs | 143 (28.3%) | 64 (32.0%) | 46 (23.0%) | 33 (31.4%) | 0.10 |
| Jaundice | 107 (21.2%) | 42 (21.0%) | 25 (12.5%) | 40 (38.1%) | <0.001 |
| Itchy skin | 91 (18.0%) | 19 (9.5%) | 40 (20.0%) | 32 (30.5%) | <0.001 |
| Pale-colored stool | 84 (16.6%) | 30 (15.0%) | 23 (11.5%) | 31 (29.5%) | <0.001 |
| Other symptoms | 44 (8.7%) | 34 (17.0%) | 3 (1.5%) | 7 (6.7%) | <0.001 |
| I don't know / I am not sure | 18 (3.6%) | 0 (0.0%) | 17 (8.5%) | 1 (1.0%) | <0.001 |
| **S6B: Current CLD symptoms (in the last month) (multiple responses possible), n (%)** | | | | | |
| Fatigue that is not related to an effort and which persists despite sleep and rest | 505 (100.0%) | 200 (100.0%) | 200 (100.0%) | 105 (100.0%) | >0.9 |
| Abdominal pain | 155 (30.7%) | 65 (32.5%) | 38 (19.0%) | 52 (49.5%) | <0.001 |
| Dark-colored urine | 120 (23.8%) | 40 (20.0%) | 48 (24.0%) | 32 (30.5%) | 0.12 |
| Nausea or vomiting | 119 (23.6%) | 57 (28.5%) | 34 (17.0%) | 28 (26.7%) | 0.018 |
| Swelling in the arms or legs | 113 (22.4%) | 56 (28.0%) | 26 (13.0%) | 31 (29.5%) | <0.001 |
| Itchy skin | 70 (13.9%) | 10 (5.0%) | 41 (20.5%) | 19 (18.1%) | <0.001 |
| Jaundice | 58 (11.5%) | 19 (9.5%) | 12 (6.0%) | 27 (25.7%) | <0.001 |
| Pale-colored stool | 57 (11.3%) | 22 (11.0%) | 11 (5.5%) | 24 (22.9%) | <0.001 |
| Other symptoms | 30 (5.9%) | 20 (10.0%) | 3 (1.5%) | 7 (6.7%) | 0.001 |
| I don't know / I am not sure | 0 (0.0%) | 0 (0.0%) | 0 (0.0%) | 0 (0.0%) | >0.9 |
| **A2.1: Number of years since the symptoms' onset (if reported symptoms at diagnosis or currently)–Fatigue** | | | | | <0.001 |
| N | 475 | 192 | 178 | 105 |  |
| Mean (SD) | 3.1 (4.9) | 1.6 (1.5) | 4.6 (6.3) | 3.4 (5.3) |  |
| Median (IQR) | 2 (1, 3) | 1 (1, 2) | 2 (1, 5) | 2 (1, 5) |  |
| Range | 0–47 | 0–8 | 0–38 | 0–47 |  |
| I do not recall (n, %) | 30 (5.9%) | 8 (4.0%) | 22 (11.0%) | 0 (0.0%) |  |
| **A2.2: Number of years since the symptoms' onset (if reported symptoms at diagnosis or currently)–Jaundice** | | | | | 0.033 |
| N | 114 | 43 | 27 | 44 |  |
| Mean (SD) | 3.2 (4.9) | 1.6 (1.5) | 5.9 (8.7) | 3.2 (3.1) |  |
| Median (IQR) | 2 (1, 3) | 1 (1, 3) | 2 (1, 9) | 2 (1, 5) |  |
| Range | 0–30 | 0–6 | 0–30 | 0–12 |  |
| I do not recall (n, %) | 5 (4.2%) | 3 (6.5%) | 1 (3.6%) | 1 (2.2%) |  |
| **A2.3: Number of years since the symptoms' onset (if reported symptoms at diagnosis or currently)–Abdominal pain** | | | | | 0.005 |
| N | 217 | 79 | 65 | 73 |  |
| Mean (SD) | 2.5 (3.2) | 1.8 (1.7) | 2.1 (2.2) | 3.6 (4.5) |  |
| Median (IQR) | 1 (1, 3) | 1 (1, 3) | 1 (1, 3) | 2 (1, 5) |  |
| Range | 0–30 | 0–7 | 0–12 | 0–30 |  |
| I do not recall (n, %) | 3 (1.4%) | 1 (1.2%) | 1 (1.5%) | 1 (1.4%) |  |
| **A2.4: Number of years since the symptoms' onset (if reported symptoms at diagnosis or currently)–Nausea or vomiting** | | | | | 0.3 |
| N | 181 | 79 | 56 | 46 |  |
| Mean (SD) | 2.9 (4.4) | 1.8 (1.6) | 4.3 (6.7) | 3.1 (3.4) |  |
| Median (IQR) | 1 (1, 3) | 1 (1, 3) | 1 (1, 5) | 2 (1, 3) |  |
| Range | 0–30 | 0–7 | 0–30 | 0–15 |  |
| I do not recall (n, %) | 7 (3.7%) | 1 (1.2%) | 5 (8.2%) | 1 (2.1%) |  |
| **A2.5: Number of years since the symptoms' onset (if reported symptoms at diagnosis or currently)–Pale-colored stool** | | | | | <0.001 |
| N | 95 | 36 | 25 | 34 |  |
| Mean (SD) | 2.8 (3.8) | 1.2 (1.4) | 4.3 (5.4) | 3.5 (3.5) |  |
| Median (IQR) | 2 (1, 3) | 1 (0, 1) | 2 (1, 4) | 2 (1, 5) |  |
| Range | 0–20 | 0–5 | 0–20 | 0–15 |  |
| I do not recall (n, %) | 4 (4.0%) | 2 (5.3%) | 2 (7.4%) | 0 (0.0%) |  |
| **A2.6: Number of years since the symptoms' onset (if reported symptoms at diagnosis or currently)–Dark-colored urine** | | | | | <0.001 |
| N | 158 | 54 | 63 | 41 |  |
| Mean (SD) | 2.8 (4.1) | 1.1 (1.3) | 3.9 (5.6) | 3.4 (3.1) |  |
| Median (IQR) | 1 (1, 4) | 1 (0, 1) | 2 (1, 4) | 3 (0, 5) |  |
| Range | 0–30 | 0–5 | 0–30 | 0–10 |  |
| I do not recall (n, %) | 9 (5.4%) | 6 (10.0%) | 3 (4.5%) | 0 (0.0%) |  |
| **A2.7: Number of years since the symptoms' onset (if reported symptoms at diagnosis or currently)–Swelling in the arms or legs** | | | | | <0.001 |
| N | 155 | 65 | 47 | 43 |  |
| Mean (SD) | 2.7 (3.2) | 1.5 (1.4) | 3.3 (3.9) | 3.9 (3.7) |  |
| Median (IQR) | 2 (1, 3) | 1 (1, 2) | 2 (1, 4) | 3 (1, 6) |  |
| Range | 0–20 | 0–7 | 0–20 | 0–15 |  |
| I do not recall (n, %) | 4 (2.5%) | 2 (3.0%) | 2 (4.1%) | 0 (0.0%) |  |
| **A2.8: Number of years since the symptoms' onset (if reported symptoms at diagnosis or currently)–Itchy skin** | | | | | 0.018 |
| N | 103 | 19 | 48 | 36 |  |
| Mean (SD) | 3.5 (5.0) | 1.5 (1.5) | 4.3 (6.5) | 3.6 (3.4) |  |
| Median (IQR) | 2 (1, 5) | 1 (1, 1) | 2 (1, 5) | 2 (1, 5) |  |
| Range | 0–30 | 0–5 | 0–30 | 0–15 |  |
| I do not recall (n, %) | 10 (8.8%) | 3 (13.6%) | 7 (12.7%) | 0 (0.0%) |  |
| **A2.9: Number of years since the symptoms' onset (if reported symptoms at diagnosis or currently)–Other** | | | | | 0.4 |
| N | 49 | 38 | 3 | 8 |  |
| Mean (SD) | 2.2 (3.2) | 1.9 (2.0) | 7.7 (10.7) | 1.8 (2.3) |  |
| Median (IQR) | 1 (1, 2) | 1 (1, 2) | 2 (2, 11) | 1 (1, 2) |  |
| Range | 0–20 | 0–8 | 1–20 | 0–7 |  |
| I do not recall (n, %) | 1 (2.0%) | 0 (0.0%) | 1 (25.0%) | 0 (0.0%) |  |
| **A3.1: Perceived severity of current symptoms–Jaundice (if reported in current CLD symptoms), n (%)** | | | | | 0.012 |
| Not at all a problem | 6 (10.3%) | 2 (10.5%) | 3 (25.0%) | 1 (3.7%) |  |
| Minor problem | 15 (25.9%) | 6 (31.6%) | 3 (25.0%) | 6 (22.2%) |  |
| Moderate problem | 23 (39.7%) | 3 (15.8%) | 3 (25.0%) | 17 (63.0%) |  |
| Serious problem | 14 (24.1%) | 8 (42.1%) | 3 (25.0%) | 3 (11.1%) |  |
| **A3.2: Perceived severity of current symptoms–Abdominal pain (if reported in current CLD symptoms), n (%)** | | | | | <0.001 |
| Not at all a problem | 1 (0.6%) | 0 (0.0%) | 0 (0.0%) | 1 (1.9%) |  |
| Minor problem | 18 (11.6%) | 1 (1.5%) | 9 (23.7%) | 8 (15.4%) |  |
| Moderate problem | 78 (50.3%) | 30 (46.2%) | 21 (55.3%) | 27 (51.9%) |  |
| Serious problem | 58 (37.4%) | 34 (52.3%) | 8 (21.1%) | 16 (30.8%) |  |
| **A3.3: Perceived severity of current symptoms–Fatigue (if reported in current CLD symptoms), n (%)** | | | | | <0.001 |
| Not at all a problem | 6 (1.2%) | 3 (1.5%) | 3 (1.5%) | 0 (0.0%) |  |
| Minor problem | 104 (20.6%) | 25 (12.5%) | 70 (35.0%) | 9 (8.6%) |  |
| Moderate problem | 259 (51.3%) | 100 (50.0%) | 106 (53.0%) | 53 (50.5%) |  |
| Serious problem | 136 (26.9%) | 72 (36.0%) | 21 (10.5%) | 43 (41.0%) |  |
| **A3.4: Perceived severity of current symptoms–Nausea or vomiting (if reported in current CLD symptoms), n (%)** | | | | | 0.10 |
| Not at all a problem | 0 (0.0%) | 0 (0.0%) | 0 (0.0%) | 0 (0.0%) |  |
| Minor problem | 27 (22.7%) | 8 (14.0%) | 8 (23.5%) | 11 (39.3%) |  |
| Moderate problem | 61 (51.3%) | 30 (52.6%) | 18 (52.9%) | 13 (46.4%) |  |
| Serious problem | 31 (26.1%) | 19 (33.3%) | 8 (23.5%) | 4 (14.3%) |  |
| **A3.5: Perceived severity of current symptoms–Pale colored-stool (if reported in current CLD symptoms), n (%)** | | | | | 0.050 |
| Not at all a problem | 5 (8.8%) | 0 (0.0%) | 1 (9.1%) | 4 (16.7%) |  |
| Minor problem | 17 (29.8%) | 6 (27.3%) | 5 (45.5%) | 6 (25.0%) |  |
| Moderate problem | 30 (52.6%) | 11 (50.0%) | 5 (45.5%) | 14 (58.3%) |  |
| Serious problem | 5 (8.8%) | 5 (22.7%) | 0 (0.0%) | 0 (0.0%) |  |
| **A3.6: Perceived severity of current symptoms–Dark colored-urine (if reported in current CLD symptoms), n (%)** | | | | | 0.003 |
| Not at all a problem | 4 (3.3%) | 1 (2.5%) | 2 (4.2%) | 1 (3.1%) |  |
| Minor problem | 37 (30.8%) | 5 (12.5%) | 20 (41.7%) | 12 (37.5%) |  |
| Moderate problem | 58 (48.3%) | 20 (50.0%) | 23 (47.9%) | 15 (46.9%) |  |
| Serious problem | 21 (17.5%) | 14 (35.0%) | 3 (6.2%) | 4 (12.5%) |  |
| **A3.7: Perceived severity of current symptoms–Swelling in the arms or legs (if reported in current CLD symptoms), n (%)** | | | | | 0.2 |
| Not at all a problem | 1 (0.9%) | 1 (1.8%) | 0 (0.0%) | 0 (0.0%) |  |
| Minor problem | 25 (22.1%) | 12 (21.4%) | 5 (19.2%) | 8 (25.8%) |  |
| Moderate problem | 57 (50.4%) | 23 (41.1%) | 15 (57.7%) | 19 (61.3%) |  |
| Serious problem | 30 (26.5%) | 20 (35.7%) | 6 (23.1%) | 4 (12.9%) |  |
| **A3.8: Perceived severity of current symptoms–Itchy skin (if reported in current CLD symptoms), n (%)** | | | | | 0.008 |
| Not at all a problem | 1 (1.4%) | 0 (0.0%) | 0 (0.0%) | 1 (5.3%) |  |
| Minor problem | 22 (31.4%) | 3 (30.0%) | 13 (31.7%) | 6 (31.6%) |  |
| Moderate problem | 38 (54.3%) | 2 (20.0%) | 26 (63.4%) | 10 (52.6%) |  |
| Serious problem | 9 (12.9%) | 5 (50.0%) | 2 (4.9%) | 2 (10.5%) |  |
| **A3.9: Perceived severity of current symptoms–Other (if reported in current CLD symptoms), n (%)** | | | | | 0.8 |
| Not at all a problem | 0 (0.0%) | 0 (0.0%) | 0 (0.0%) | 0 (0.0%) |  |
| Minor problem | 5 (16.7%) | 3 (15.0%) | 0 (0.0%) | 2 (28.6%) |  |
| Moderate problem | 22 (73.3%) | 14 (70.0%) | 3 (100.0%) | 5 (71.4%) |  |
| Serious problem | 3 (10.0%) | 3 (15.0%) | 0 (0.0%) | 0 (0.0%) |  |
| **S8: Cirrhosis Diagnosis, n (%)** | | | | | <0.001 |
| Yes | 46 (9.1%) | 1 (0.5%) | 31 (15.5%) | 14 (13.3%) |  |
| No | 454 (89.9%) | 198 (99.0%) | 166 (83.0%) | 90 (85.7%) |  |
| I do not know | 5 (1.0%) | 1 (0.5%) | 3 (1.5%) | 1 (1.0%) |  |
| **S9.1: Symptoms that led to seek medical attention (if diagnosis of Cirrhosis or don't know), Haematemesis: at least one episode of vomiting blood, n (%)** | 0 (0.0%) | 0 (0.0%) | 0 (0.0%) | 0 (0.0%) | >0.9 |
| **S9.2: Symptoms that led to seek medical attention (if diagnosis of Cirrhosis or don't know), Ascites: fluid accumulation in the abdomen that required drainage, n (%)** | 0 (0.0%) | 0 (0.0%) | 0 (0.0%) | 0 (0.0%) | >0.9 |
| **S9.3: Symptoms that led to seek medical attention (if diagnosis of Cirrhosis or don't know), Encephalopathy: confusion and/or altered consciousness and/or forgetfulness and/or changed in mood/personality, n (%)** | 0 (0.0%) | 0 (0.0%) | 0 (0.0%) | 0 (0.0%) | >0.9 |
| **S7: Treatment of fatigue, n (%)** |  |  |  |  | >0.9 |
| Yes | 0 (0.0%) | 0 (0.0%) | 0 (0.0%) | 0 (0.0%) |  |
| No | 505 (100.0%) | 200 (100.0%) | 200 (100.0%) | 105 (100.0%) |  |
| **A4.1: Physical fatigue characterization, n (%)** | | | | | 0.019 |
| Yes | 482 (95.4%) | 189 (94.5%) | 188 (94.0%) | 105 (100.0%) |  |
| No | 23 (4.6%) | 11 (5.5%) | 12 (6.0%) | 0 (0.0%) |  |
| **A4.2: Mental fatigue characterization, n (%)** | | | | | <0.001 |
| Yes | 356 (70.5%) | 111 (55.5%) | 167 (83.5%) | 78 (74.3%) |  |
| No | 149 (29.5%) | 89 (44.5%) | 33 (16.5%) | 27 (25.7%) |  |
| **A5: Fatigue frequency, n (%)** | | | | | <0.001 |
| Every day or almost every day | 169 (33.5%) | 61 (30.5%) | 42 (21.0%) | 66 (62.9%) |  |
| 2 to 5 days per week | 244 (48.3%) | 105 (52.5%) | 106 (53.0%) | 33 (31.4%) |  |
| 1 day per week | 74 (14.7%) | 27 (13.5%) | 41 (20.5%) | 6 (5.7%) |  |
| Less often | 18 (3.6%) | 7 (3.5%) | 11 (5.5%) | 0 (0.0%) |  |
| **A6: Discuss fatigue with a doctor, n (%)** | | | | | <0.001 |
| Yes | 320 (63.4%) | 82 (41.0%) | 149 (74.5%) | 89 (84.8%) |  |
| No | 185 (36.6%) | 118 (59.0%) | 51 (25.5%) | 16 (15.2%) |  |
| *^1^* Fisher’s exact test; Pearson’s Chi-squared test; Kruskal-Wallis rank sum test; Chi-Squared Test with Monte Carlo Simulation | | | | | |

AIDS, acquired immune deficiency syndrome; CKD, chronic kidney disease; CLD, chronic liver disease; HIV, human immunodeficiency virus; IQR, interquartile range; MASH, metabolic dysfunction-associated steatohepatitis; MASLD, metabolic dysfunction-associated steatotic liver disease; NASH, non-alcoholic steatohepatitis; NAFLD, non-alcoholic fatty liver disease; PROMIS-29+2, Patient-Reported Outcomes Measurement Information System; SD, standard deviation.

**Supplementary Table 4**. Treatment data

| **Base: All participants** | **N = 505** | **Country** | | | **p-value***^1^* |
| --- | --- | --- | --- | --- | --- |
|  |  | **India, N = 200** | **China, N = 200** | **Mexico, N = 105** |  |
| **A7: CLD treatments (multiple responses possible), n (%)** |  |  |  |  |  |
| Antioxidants | 93 (18.4%) | 15 (7.5%) | 53 (26.5%) | 25 (23.8%) | <0.001 |
| Statins, a medicine that lowers the level of cholesterol or lipids/fats in the blood | 75 (14.9%) | 17 (8.5%) | 39 (19.5%) | 19 (18.1%) | 0.005 |
| Antiviral drugs | 71 (14.1%) | 3 (1.5%) | 58 (29.0%) | 10 (9.5%) | <0.001 |
| A medicine that improves blood sugar control | 29 (5.7%) | 8 (4.0%) | 10 (5.0%) | 11 (10.5%) | <0.001 |
| A medicine to help reduce my consumption of alcohol | 9 (1.8%) | 1 (0.5%) | 7 (3.5%) | 1 (1.0%) | <0.001 |
| Pentoxifylline, a medicine to help improve blood flow and reduce inflammation in the liver | 9 (1.8%) | 3 (1.5%) | 3 (1.5%) | 3 (2.9%) | <0.001 |
| Diuretic | 9 (1.8%) | 0 (0.0%) | 6 (3.0%) | 3 (2.9%) | <0.001 |
| Ursodeoxycholic acid, a medicine that helps in dissolving gallstones and improving bile flow | 7 (1.4%) | 1 (0.5%) | 6 (3.0%) | 0 (0.0%) | <0.001 |
| Calcium and Vitamin D supplementation | 3 (0.6%) | 1 (0.5%) | 1 (0.5%) | 1 (1.0%) | 0.076 |
| Beta-blocker | 3 (0.6%) | 0 (0.0%) | 2 (1.0%) | 1 (1.0%) | <0.001 |
| Cholestyramine, a medicine that helps lower the levels of bile acids in the body | 2 (0.4%) | 1 (0.5%) | 1 (0.5%) | 0 (0.0%) | 0.14 |
| Interferon injections, a medicine that is injected to help the body's immune system fight viral infections | 2 (0.4%) | 0 (0.0%) | 2 (1.0%) | 0 (0.0%) | 0.035 |
| Obeticholic acid, a medicine that helps increase the flow of bile acids out of the liver and reduces exposure to toxic bile acids | 1 (0.2%) | 0 (0.0%) | 0 (0.0%) | 1 (1.0%) | 0.038 |
| Immunosuppressant | 1 (0.2%) | 0 (0.0%) | 1 (0.5%) | 0 (0.0%) | 0.3 |
| Corticosteroid, or medicines that help to reduce inflammation in the body | 1 (0.2%) | 1 (0.5%) | 0 (0.0%) | 0 (0.0%) | 0.038 |
| Adalimumab, a medicine that targets specific proteins in the body that are causing inflammation in the liver | 1 (0.2%) | 0 (0.0%) | 0 (0.0%) | 1 (1.0%) | 0.038 |
| Chelating agents for people with Haemochromatosis | 1 (0.2%) | 1 (0.5%) | 0 (0.0%) | 0 (0.0%) | 0.7 |
| Rifaximin | 1 (0.2%) | 0 (0.0%) | 0 (0.0%) | 1 (1.0%) | <0.001 |
| Diphenhydramine, a drug commonly prescribed to relieve symptoms of allergies and that can help relieve itching caused by certain types of chronic liver disease | 0 (0.0%) | 0 (0.0%) | 0 (0.0%) | 0 (0.0%) | 0.14 |
| Chelating agents for people with Wilson's disease | 0 (0.0%) | 0 (0.0%) | 0 (0.0%) | 0 (0.0%) | 0.4 |
| Other | 27 (5.3%) | 7 (3.5%) | 16 (8.0%) | 4 (3.8%) | 0.10 |
| I'm taking a treatment prescribed by a doctor, but I don't know what it is called/named | 64 (12.7%) | 44 (22.0%) | 16 (8.0%) | 4 (3.8%) | <0.001 |
| I'm not taking any treatment prescribed by a doctor for my liver disease | 222 (44.0%) | 121 (60.5%) | 51 (25.5%) | 50 (47.6%) | <0.001 |
| **A8: Ever received treatment prescribed by a physician for CLD (if not currently treated), n (%)** | 31 (14.0%) | 21 (17.4%) | 8 (15.7%) | 2 (4.0%) | 0.067 |
| **Ever treated for CLD, n (%)** | 314 (62.2%) | 100 (50.0%) | 157 (78.5%) | 57 (54.3%) | <0.001 |
| **A9: Treatment satisfaction (if currently receiving a treatment for CLD), n (%)** |  |  |  |  | 0.005 |
| Very satisfied | 23 (8.1%) | 9 (11.4%) | 14 (9.4%) | 0 (0.0%) |  |
| Satisfied | 138 (48.8%) | 49 (62.0%) | 63 (42.3%) | 26 (47.3%) |  |
| Neither satisfied nor dissatisfied | 103 (36.4%) | 16 (20.3%) | 60 (40.3%) | 27 (49.1%) |  |
| Dissatisfied | 18 (6.4%) | 5 (6.3%) | 11 (7.4%) | 2 (3.6%) |  |
| Very dissatisfied | 1 (0.4%) | 0 (0.0%) | 1 (0.7%) | 0 (0.0%) |  |
| **A10: Perception of treatment effectiveness (if currently receiving a treatment for CLD), n (%)** |  |  |  |  | <0.001 |
| Very good | 19 (6.7%) | 10 (12.7%) | 9 (6.0%) | 0 (0.0%) |  |
| Good | 91 (32.2%) | 40 (50.6%) | 42 (28.2%) | 9 (16.4%) |  |
| Acceptable | 161 (56.9%) | 26 (32.9%) | 91 (61.1%) | 44 (80.0%) |  |
| Poor | 12 (4.2%) | 3 (3.8%) | 7 (4.7%) | 2 (3.6%) |  |
| Very poor | 0 (0.0%) | 0 (0.0%) | 0 (0.0%) | 0 (0.0%) |  |
| **A11.1: Involvement in treatment choice for current or last CLD treatment (if ever treated), n (%)** |  |  |  |  | <0.001 |
| True | 197 (62.7%) | 38 (38.0%) | 133 (84.7%) | 26 (45.6%) |  |
| False | 117 (37.3%) | 62 (62.0%) | 24 (15.3%) | 31 (54.4%) |  |
| **A11.2: Personal preference considered in CLD treatment prescription (if ever treated), n (%)** |  |  |  |  | <0.001 |
| True | 267 (85.0%) | 74 (74.0%) | 146 (93.0%) | 47 (82.5%) |  |
| False | 47 (15.0%) | 26 (26.0%) | 11 (7.0%) | 10 (17.5%) |  |
| **A13.1: Received all information needed to fully understand treatment recommendation (if ever treated), n (%)** |  |  |  |  | <0.001 |
| Strongly disagree | 5 (1.6%) | 3 (3.0%) | 0 (0.0%) | 2 (3.5%) |  |
| Disagree | 34 (10.8%) | 11 (11.0%) | 17 (10.8%) | 6 (10.5%) |  |
| Agree | 211 (67.2%) | 56 (56.0%) | 122 (77.7%) | 33 (57.9%) |  |
| Strongly agree | 64 (20.4%) | 30 (30.0%) | 18 (11.5%) | 16 (28.1%) |  |
| **A13.2: Received enough information to feel confident that right treatment received (if ever treated), n (%)** |  |  |  |  | 0.7 |
| Strongly disagree | 1 (0.3%) | 0 (0.0%) | 1 (0.6%) | 0 (0.0%) |  |
| Disagree | 28 (8.9%) | 11 (11.0%) | 10 (6.4%) | 7 (12.3%) |  |
| Agree | 200 (63.7%) | 64 (64.0%) | 102 (65.0%) | 34 (59.6%) |  |
| Strongly agree | 85 (27.1%) | 25 (25.0%) | 44 (28.0%) | 16 (28.1%) |  |
| **A13.3: Given enough time and explanation to understand information and treatment recommendation (if ever treated), n (%)** |  |  |  |  | 0.026 |
| Strongly disagree | 8 (2.5%) | 5 (5.0%) | 1 (0.6%) | 2 (3.5%) |  |
| Disagree | 61 (19.4%) | 26 (26.0%) | 25 (15.9%) | 10 (17.5%) |  |
| Agree | 179 (57.0%) | 46 (46.0%) | 103 (65.6%) | 30 (52.6%) |  |
| Strongly agree | 66 (21.0%) | 23 (23.0%) | 28 (17.8%) | 15 (26.3%) |  |
| **A13.4: Given the needed information but did not understand it (if ever treated), n (%)** |  |  |  |  | 0.002 |
| Strongly disagree | 80 (25.5%) | 23 (23.0%) | 32 (20.4%) | 25 (43.9%) |  |
| Disagree | 152 (48.4%) | 47 (47.0%) | 83 (52.9%) | 22 (38.6%) |  |
| Agree | 61 (19.4%) | 21 (21.0%) | 36 (22.9%) | 4 (7.0%) |  |
| Strongly agree | 21 (6.7%) | 9 (9.0%) | 6 (3.8%) | 6 (10.5%) |  |
| **A14: Topics of CLD patients discussed with physicians before CLD treatment (if ever treated–multiple responses possible), n (%)** |  |  |  |  |  |
| Whether symptoms will improve | 213 (67.8%) | 61 (61.0%) | 116 (73.9%) | 36 (63.2%) | 0.069 |
| Efficacy of treatments | 200 (63.7%) | 47 (47.0%) | 117 (74.5%) | 36 (63.2%) | <0.001 |
| Treatment options | 197 (62.7%) | 53 (53.0%) | 114 (72.6%) | 30 (52.6%) | 0.001 |
| Whether quality of life will improve with treatment | 190 (60.5%) | 52 (52.0%) | 94 (59.9%) | 44 (77.2%) | 0.008 |
| Treatment side effects | 186 (59.2%) | 47 (47.0%) | 102 (65.0%) | 37 (64.9%) | 0.011 |
| Disease prognosis | 183 (58.3%) | 45 (45.0%) | 102 (65.0%) | 36 (63.2%) | 0.005 |
| The costs of the treatment and/or out of pocket expenses | 166 (52.9%) | 45 (45.0%) | 83 (52.9%) | 38 (66.7%) | 0.033 |
| None of these | 10 (3.2%) | 5 (5.0%) | 5 (3.2%) | 0 (0.0%) | 0.3 |
| **A15: Features that would most positively influence and contribute to the acceptance of a new treatment for your liver disease (ranking–if ever treated)** | | | | | |
| **A15.1: Prevent progression/worsening of fibrosis/cirrhosis** |  |  |  |  | <0.001 |
| 1 (most important) | 92 (29.3%) | 9 (9.0%) | 67 (42.7%) | 16 (28.1%) |  |
| 2 | 61 (19.4%) | 12 (12.0%) | 37 (23.6%) | 12 (21.1%) |  |
| 3 | 34 (10.8%) | 9 (9.0%) | 16 (10.2%) | 9 (15.8%) |  |
| 4 | 24 (7.6%) | 14 (14.0%) | 6 (3.8%) | 4 (7.0%) |  |
| 5 | 29 (9.2%) | 12 (12.0%) | 9 (5.7%) | 8 (14.0%) |  |
| 6 | 21 (6.7%) | 12 (12.0%) | 7 (4.5%) | 2 (3.5%) |  |
| 7 | 29 (9.2%) | 14 (14.0%) | 10 (6.4%) | 5 (8.8%) |  |
| 8 (least important) | 24 (7.6%) | 18 (18.0%) | 5 (3.2%) | 1 (1.8%) |  |
| N | 314 | 100 | 157 | 57 | <0.001 |
| Mean (SD) | 3.4 (2.4) | 4.9 (2.3) | 2.6 (2.1) | 3.1 (2.0) |  |
| Median (IQR) | 3 (1, 5) | 5 (3, 7) | 2 (1, 3) | 3 (1, 5) |  |
| Range | 1–8 | 1–8 | 1–8 | 1–8 |  |
| **A15.2: Prevent fatigue** |  |  |  |  | <0.001 |
| 1 (most important) | 78 (24.8%) | 29 (29.0%) | 27 (17.2%) | 22 (38.6%) |  |
| 2 | 45 (14.3%) | 11 (11.0%) | 19 (12.1%) | 15 (26.3%) |  |
| 3 | 33 (10.5%) | 6 (6.0%) | 19 (12.1%) | 8 (14.0%) |  |
| 4 | 38 (12.1%) | 10 (10.0%) | 24 (15.3%) | 4 (7.0%) |  |
| 5 | 37 (11.8%) | 8 (8.0%) | 26 (16.6%) | 3 (5.3%) |  |
| 6 | 22 (7.0%) | 6 (6.0%) | 14 (8.9%) | 2 (3.5%) |  |
| 7 | 30 (9.6%) | 10 (10.0%) | 20 (12.7%) | 0 (0.0%) |  |
| 8 (least important) | 31 (9.9%) | 20 (20.0%) | 8 (5.1%) | 3 (5.3%) |  |
| N | 314 | 100 | 157 | 57 | <0.001 |
| Mean (SD) | 3.8 (2.4) | 4.2 (2.8) | 4.1 (2.2) | 2.5 (1.9) |  |
| Median (IQR) | 4 (2, 6) | 4 (1, 7) | 4 (2, 6) | 2 (1, 3) |  |
| Range | 1–8 | 1–8 | 1–8 | 1–8 |  |
| **A15.3: Limited or manageable side-effects** |  |  |  |  | 0.082 |
| 1 (most important) | 32 (10.2%) | 12 (12.0%) | 14 (8.9%) | 6 (10.5%) |  |
| 2 | 51 (16.2%) | 10 (10.0%) | 32 (20.4%) | 9 (15.8%) |  |
| 3 | 71 (22.6%) | 16 (16.0%) | 39 (24.8%) | 16 (28.1%) |  |
| 4 | 49 (15.6%) | 14 (14.0%) | 27 (17.2%) | 8 (14.0%) |  |
| 5 | 35 (11.1%) | 12 (12.0%) | 15 (9.6%) | 8 (14.0%) |  |
| 6 | 41 (13.1%) | 16 (16.0%) | 17 (10.8%) | 8 (14.0%) |  |
| 7 | 25 (8.0%) | 14 (14.0%) | 9 (5.7%) | 2 (3.5%) |  |
| 8 (least important) | 10 (3.2%) | 6 (6.0%) | 4 (2.5%) | 0 (0.0%) |  |
| N | 314 | 100 | 157 | 57 | 0.013 |
| Mean (SD) | 3.9 (1.9) | 4.4 (2.1) | 3.7 (1.8) | 3.6 (1.7) |  |
| Median (IQR) | 4 (2, 5) | 4 (3, 6) | 3 (2, 5) | 3 (2, 5) |  |
| Range | 1–8 | 1–8 | 1–8 | 1–7 |  |
| **A15.4: Convenient dosing schedule** |  |  |  |  | <0.001 |
| 1 (most important) | 9 (2.9%) | 4 (4.0%) | 5 (3.2%) | 0 (0.0%) |  |
| 2 | 20 (6.4%) | 10 (10.0%) | 9 (5.7%) | 1 (1.8%) |  |
| 3 | 36 (11.5%) | 15 (15.0%) | 19 (12.1%) | 2 (3.5%) |  |
| 4 | 48 (15.3%) | 15 (15.0%) | 27 (17.2%) | 6 (10.5%) |  |
| 5 | 56 (17.8%) | 20 (20.0%) | 31 (19.7%) | 5 (8.8%) |  |
| 6 | 56 (17.8%) | 8 (8.0%) | 38 (24.2%) | 10 (17.5%) |  |
| 7 | 45 (14.3%) | 13 (13.0%) | 14 (8.9%) | 18 (31.6%) |  |
| 8 (least important) | 44 (14.0%) | 15 (15.0%) | 14 (8.9%) | 15 (26.3%) |  |
| N | 314 | 100 | 157 | 57 | <0.001 |
| Mean (SD) | 5.2 (1.9) | 4.9 (2.1) | 5.0 (1.8) | 6.4 (1.5) |  |
| Median (IQR) | 5 (4, 7) | 5 (3, 7) | 5 (4, 6) | 7 (6, 8) |  |
| Range | 1–8 | 1–8 | 1–8 | 2–8 |  |
| **A15.5: How the treatment is taken** |  |  |  |  | 0.053 |
| 1 (most important) | 31 (9.9%) | 7 (7.0%) | 21 (13.4%) | 3 (5.3%) |  |
| 2 | 35 (11.1%) | 16 (16.0%) | 17 (10.8%) | 2 (3.5%) |  |
| 3 | 54 (17.2%) | 13 (13.0%) | 32 (20.4%) | 9 (15.8%) |  |
| 4 | 36 (11.5%) | 11 (11.0%) | 20 (12.7%) | 5 (8.8%) |  |
| 5 | 49 (15.6%) | 14 (14.0%) | 27 (17.2%) | 8 (14.0%) |  |
| 6 | 42 (13.4%) | 13 (13.0%) | 16 (10.2%) | 13 (22.8%) |  |
| 7 | 41 (13.1%) | 17 (17.0%) | 14 (8.9%) | 10 (17.5%) |  |
| 8 (least important) | 26 (8.3%) | 9 (9.0%) | 10 (6.4%) | 7 (12.3%) |  |
| N | 314 | 100 | 157 | 57 | 0.001 |
| Mean (SD) | 4.5 (2.1) | 4.6 (2.2) | 4.1 (2.1) | 5.2 (2.0) |  |
| Median (IQR) | 5 (3, 6) | 5 (3, 7) | 4 (3, 6) | 6 (4, 7) |  |
| Range | 1–8 | 1–8 | 1–8 | 1–8 |  |
| **A15.6: How often you have to take your medication** |  |  |  |  | <0.001 |
| 1 (most important) | 13 (4.1%) | 10 (10.0%) | 3 (1.9%) | 0 (0.0%) |  |
| 2 | 35 (11.1%) | 12 (12.0%) | 20 (12.7%) | 3 (5.3%) |  |
| 3 | 25 (8.0%) | 15 (15.0%) | 8 (5.1%) | 2 (3.5%) |  |
| 4 | 41 (13.1%) | 10 (10.0%) | 23 (14.6%) | 8 (14.0%) |  |
| 5 | 51 (16.2%) | 19 (19.0%) | 19 (12.1%) | 13 (22.8%) |  |
| 6 | 59 (18.8%) | 12 (12.0%) | 29 (18.5%) | 18 (31.6%) |  |
| 7 | 53 (16.9%) | 11 (11.0%) | 34 (21.7%) | 8 (14.0%) |  |
| 8 (least important) | 37 (11.8%) | 11 (11.0%) | 21 (13.4%) | 5 (8.8%) |  |
| N | 314 | 100 | 157 | 57 | 0.004 |
| Mean (SD) | 5.1 (2.0) | 4.5 (2.2) | 5.3 (2.0) | 5.5 (1.5) |  |
| Median (IQR) | 5 (4, 7) | 5 (3, 6) | 6 (4, 7) | 6 (5, 6) |  |
| Range | 1–8 | 1–8 | 1–8 | 2–8 |  |
| **A15.7: A treatment that is financially accessible to me** |  |  |  |  | 0.022 |
| 1 (most important) | 37 (11.8%) | 13 (13.0%) | 18 (11.5%) | 6 (10.5%) |  |
| 2 | 48 (15.3%) | 19 (19.0%) | 18 (11.5%) | 11 (19.3%) |  |
| 3 | 38 (12.1%) | 16 (16.0%) | 14 (8.9%) | 8 (14.0%) |  |
| 4 | 47 (15.0%) | 10 (10.0%) | 21 (13.4%) | 16 (28.1%) |  |
| 5 | 30 (9.6%) | 9 (9.0%) | 15 (9.6%) | 6 (10.5%) |  |
| 6 | 38 (12.1%) | 13 (13.0%) | 23 (14.6%) | 2 (3.5%) |  |
| 7 | 30 (9.6%) | 10 (10.0%) | 16 (10.2%) | 4 (7.0%) |  |
| 8 (least important) | 46 (14.6%) | 10 (10.0%) | 32 (20.4%) | 4 (7.0%) |  |
| N | 314 | 100 | 157 | 57 | 0.006 |
| Mean (SD) | 4.4 (2.3) | 4.1 (2.3) | 4.8 (2.4) | 3.8 (2.0) |  |
| Median (IQR) | 4 (2, 6) | 4 (2, 6) | 5 (3, 7) | 4 (2, 5) |  |
| Range | 1–8 | 1–8 | 1–8 | 1–8 |  |
| **A15.8: A treatment that does not put me at risk of disclosing my status to others** |  |  |  |  | <0.001 |
| 1 (most important) | 22 (7.0%) | 16 (16.0%) | 2 (1.3%) | 4 (7.0%) |  |
| 2 | 19 (6.1%) | 10 (10.0%) | 5 (3.2%) | 4 (7.0%) |  |
| 3 | 23 (7.3%) | 10 (10.0%) | 10 (6.4%) | 3 (5.3%) |  |
| 4 | 31 (9.9%) | 16 (16.0%) | 9 (5.7%) | 6 (10.5%) |  |
| 5 | 27 (8.6%) | 6 (6.0%) | 15 (9.6%) | 6 (10.5%) |  |
| 6 | 35 (11.1%) | 20 (20.0%) | 13 (8.3%) | 2 (3.5%) |  |
| 7 | 61 (19.4%) | 11 (11.0%) | 40 (25.5%) | 10 (17.5%) |  |
| 8 (least important) | 96 (30.6%) | 11 (11.0%) | 63 (40.1%) | 22 (38.6%) |  |
| N | 314 | 100 | 157 | 57 | <0.001 |
| Mean (SD) | 5.7 (2.3) | 4.5 (2.3) | 6.5 (1.8) | 5.8 (2.4) |  |
| Median (IQR) | 7 (4, 8) | 4 (2, 6) | 7 (5, 8) | 7 (4, 8) |  |
| Range | 1–8 | 1–8 | 1–8 | 1–8 |  |
| **A16: Lifestyle changes to be implemented to manage CLD (multiple responses possible), n (%)** |  |  |  |  |  |
| Nutritional adjustments or dietary restrictions | 364 (72.1%) | 121 (60.5%) | 164 (82.0%) | 79 (75.2%) | <0.001 |
| Being physically active / regularly exercise | 338 (66.9%) | 143 (71.5%) | 142 (71.0%) | 53 (50.5%) | <0.001 |
| Reducing salt intake | 259 (51.3%) | 88 (44.0%) | 102 (51.0%) | 69 (65.7%) | 0.001 |
| Weight loss | 236 (46.7%) | 99 (49.5%) | 95 (47.5%) | 42 (40.0%) | 0.3 |
| Stopping alcohol consumption | 222 (44.0%) | 39 (19.5%) | 120 (60.0%) | 63 (60.0%) | <0.001 |
| Drinking less fluids | 92 (18.2%) | 66 (33.0%) | 21 (10.5%) | 5 (4.8%) | <0.001 |
| Other | 19 (3.8%) | 16 (8.0%) | 1 (0.5%) | 2 (1.9%) | <0.001 |
| None of the above | 10 (2.0%) | 4 (2.0%) | 1 (0.5%) | 5 (4.8%) | 0.046 |
| **A17: Non-prescription fatigue treatments currently taken (multiple responses possible), n (%)** |  |  |  |  |  |
| Lifestyle or behavioural modification therapies | 202 (40.0%) | 82 (41.0%) | 107 (53.5%) | 13 (12.4%) | <0.001 |
| Vitamins | 182 (36.0%) | 41 (20.5%) | 101 (50.5%) | 40 (38.1%) | <0.001 |
| Traditional medicine such as herbal remedies, traditional Chinese medicine, naturopathy, traditional Indian medicine or ayurvedic therapy | 98 (19.4%) | 37 (18.5%) | 47 (23.5%) | 14 (13.3%) | 0.094 |
| Acupuncture | 36 (7.1%) | 15 (7.5%) | 17 (8.5%) | 4 (3.8%) | 0.3 |
| Other herbal or traditional remedies not listed as examples above | 34 (6.7%) | 25 (12.5%) | 3 (1.5%) | 6 (5.7%) | <0.001 |
| Other | 37 (7.3%) | 27 (13.5%) | 9 (4.5%) | 1 (1.0%) | <0.001 |
| None of the above | 159 (31.5%) | 60 (30.0%) | 46 (23.0%) | 53 (50.5%) | <0.001 |
| *^1^* Pearson’s Chi-squared test; Fisher’s exact test; Kruskal-Wallis rank sum test; Chi-Squared Test with Monte Carlo Simulation | | | | | |

CLD, chronic liver disease; IQR, interquartile range; SD, standard deviation.

**Supplementary Table 5**. Socio-behavioural characteristics

| **Base: All participants** | **N = 505** | **Country** | | | **p-value***^1^* |
| --- | --- | --- | --- | --- | --- |
|  |  | **India, N = 200** | **China, N = 200** | **Mexico, N = 105** |  |
| **D4: Smoking status, n (%)** |  |  |  |  | <0.001 |
| Yes, I’m currently smoking | 59 (11.7%) | 15 (7.5%) | 32 (16.0%) | 12 (11.4%) |  |
| No, I quit smoking | 137 (27.1%) | 33 (16.5%) | 52 (26.0%) | 52 (49.5%) |  |
| No, I never smoked | 309 (61.2%) | 152 (76.0%) | 116 (58.0%) | 41 (39.0%) |  |
| **D5: Smoking habit per month (past 30 days) if currently smoking, n (%)** |  |  |  |  | 0.026 |
| More than 20 days | 35 (59.3%) | 11 (73.3%) | 20 (62.5%) | 4 (33.3%) |  |
| About 10 to 20 days | 14 (23.7%) | 1 (6.7%) | 9 (28.1%) | 4 (33.3%) |  |
| About 1 to 9 days | 8 (13.6%) | 3 (20.0%) | 1 (3.1%) | 4 (33.3%) |  |
| No days | 2 (3.4%) | 0 (0.0%) | 2 (6.2%) | 0 (0.0%) |  |
| **D6: Use of tobacco or other nicotine products (multiple responses possible), n (%)** |  |  |  |  |  |
| Other tobacco products besides cigarettes, cigars, pipes or vapes | 29 (5.7%) | 9 (4.5%) | 9 (4.5%) | 11 (10.5%) | 0.064 |
| Nicotine replacement products such as gum or patch | 26 (5.1%) | 13 (6.5%) | 8 (4.0%) | 5 (4.8%) | 0.5 |
| Chewing tobacco or snuff | 18 (3.6%) | 11 (5.5%) | 7 (3.5%) | 0 (0.0%) | 0.035 |
| None of the above | 439 (86.9%) | 169 (84.5%) | 181 (90.5%) | 89 (84.8%) | 0.2 |
| **D7: Use of marijuana (for non-medical purposes) in the past 12 months, n (%)** |  |  |  |  | <0.001 |
| Yes, monthly or more often | 1 (0.3%) | 1 (0.5%) | 0 (NA) | 0 (0.0%) |  |
| Yes, every 2 to 6 months | 25 (8.2%) | 23 (11.5%) | 0 (NA) | 2 (1.9%) |  |
| Yes, less often | 18 (5.9%) | 18 (9.0%) | 0 (NA) | 0 (0.0%) |  |
| No | 253 (83.0%) | 151 (75.5%) | 0 (NA) | 102 (97.1%) |  |
| Prefer not to answer | 8 (2.6%) | 7 (3.5%) | 0 (NA) | 1 (1.0%) |  |
| **D8: Frequency of exercise, n (%)** |  |  |  |  | <0.001 |
| 1–Never | 111 (22.0%) | 36 (18.0%) | 35 (17.5%) | 40 (38.1%) |  |
| 2 | 139 (27.5%) | 24 (12.0%) | 79 (39.5%) | 36 (34.3%) |  |
| 3 | 117 (23.2%) | 33 (16.5%) | 60 (30.0%) | 24 (22.9%) |  |
| 4 | 58 (11.5%) | 42 (21.0%) | 14 (7.0%) | 2 (1.9%) |  |
| 5 times/week or more | 80 (15.8%) | 65 (32.5%) | 12 (6.0%) | 3 (2.9%) |  |
| **D9: Level of effort exerted when exercising, n (%)** |  |  |  |  | <0.001 |
| 1–None or very low | 4 (1.0%) | 1 (0.6%) | 2 (1.2%) | 1 (1.5%) |  |
| 2 | 28 (7.1%) | 7 (4.3%) | 14 (8.5%) | 7 (10.8%) |  |
| 3 | 52 (13.2%) | 8 (4.9%) | 35 (21.2%) | 9 (13.8%) |  |
| 4 | 56 (14.2%) | 17 (10.4%) | 33 (20.0%) | 6 (9.2%) |  |
| 5 | 62 (15.7%) | 13 (7.9%) | 40 (24.2%) | 9 (13.8%) |  |
| 6 | 39 (9.9%) | 11 (6.7%) | 21 (12.7%) | 7 (10.8%) |  |
| 7 | 49 (12.4%) | 30 (18.3%) | 13 (7.9%) | 6 (9.2%) |  |
| 8 | 65 (16.5%) | 44 (26.8%) | 6 (3.6%) | 15 (23.1%) |  |
| 9 | 21 (5.3%) | 19 (11.6%) | 0 (0.0%) | 2 (3.1%) |  |
| 10–Very high | 18 (4.6%) | 14 (8.5%) | 1 (0.6%) | 3 (4.6%) |  |
| *^1^* Pearson’s Chi-squared test; Fisher’s exact test; Chi-Squared Test with Monte Carlo Simulation. | | | | | |

**Supplementary Table 6**. General health data

| **Base: All participants** | **N = 505** | **Country** | | | **p-value***^1^* |
| --- | --- | --- | --- | --- | --- |
|  |  | **India, N = 200** | **China, N = 200** | **Mexico, N = 105** |  |
| **E1: BMI** |  |  |  |  | <0.001 |
| N | 483 | 179 | 199 | 105 |  |
| Mean (SD) | 26.3 (5.7) | 25.5 (4.7) | 26.0 (6.5) | 28.3 (5.0) |  |
| Median (IQR) | 25.4 (22.5, 29.0) | 25.0 (22.1, 28.4) | 24.8 (22.5, 27.8) | 28.4 (24.4, 31.3) |  |
| Range | 14.9–58.8 | 15.6–44.6 | 14.9–58.8 | 18.0–41.5 |  |
| I don't know / prefer not to answer (n, %) | 21 (4.2%) | 20 (10.0%) | 1 (0.5%) | 0 (0.0%) |  |
| Erroneous data* | 1 (0.2%) | 1 (0.5%) | 0 (0.0%) | 0 (0.0%) |  |
| **BMI category (on those who provided height and weight), n (%)** |  |  |  |  | <0.001 |
| Underweight (BMI<18.5) | 11 (2.3%) | 4 (2.2%) | 6 (3.0%) | 1 (1.0%) |  |
| Normal weight (18.5<=BMI<25) | 217 (44.9%) | 88 (49.2%) | 96 (48.2%) | 33 (31.4%) |  |
| Overweight (25<=BMI<30) | 159 (32.9%) | 57 (31.8%) | 70 (35.2%) | 32 (30.5%) |  |
| Obesity (BMI>=30) | 96 (19.9%) | 30 (16.8%) | 27 (13.6%) | 39 (37.1%) |  |
| **E3: Current treatment (multiple responses possible), n (%)** |  |  |  |  |  |
| Antiemetics | 57 (11.3%) | 39 (19.5%) | 6 (3.0%) | 12 (11.4%) | <0.001 |
| Antihistamines | 28 (5.5%) | 12 (6.0%) | 9 (4.5%) | 7 (6.7%) | 0.7 |
| Anxiolytics | 30 (5.9%) | 8 (4.0%) | 10 (5.0%) | 12 (11.4%) | 0.026 |
| Antidepressants | 28 (5.5%) | 11 (5.5%) | 5 (2.5%) | 12 (11.4%) | 0.005 |
| Cancer treatments | 5 (1.0%) | 0 (0.0%) | 5 (2.5%) | 0 (0.0%) | 0.003 |
| Opioids | 58 (11.5%) | 20 (10.0%) | 25 (12.5%) | 13 (12.4%) | 0.7 |
| Lipid lowering agents | 93 (18.4%) | 17 (8.5%) | 57 (28.5%) | 19 (18.1%) | <0.001 |
| None of these | 328 (65.0%) | 142 (71.0%) | 123 (61.5%) | 63 (60.0%) | 0.067 |
| *^1^* Kruskal-Wallis rank sum test; Fisher’s exact test; Pearson’s Chi-squared test; Chi-Squared Test with Monte Carlo Simulation | | | | | |
| *Data were removed due to inconsistencies between height and weight.  BMI, body mass index; IQR, interquartile range; SD, standard deviation. | | | | |  |

**Supplementary Table 7**. Presence of fatigue by participant subgroup

|  | **Physical fatigue characterization, n (%)** | | **Mental fatigue characterization, n (%)** | |
| --- | --- | --- | --- | --- |
|  | ***Yes*** | ***No*** | ***Yes*** | ***No*** |
| **CLD duration** | | | | |
| Overall (N = 494) | 472 (95.5) | 22 (4.5) | 348 (70.4) | 146 (29.6) |
| <1 year (N=84) | 81 (96.4) | 3 (3.6) | 54 (64.3) | 30 (35.7) |
| 1–<2 years (N=113) | 106 (93.8) | 7 (6.2) | 80 (70.8) | 33 (29.2) |
| 2–<4 years (N=156) | 149 (95.5) | 7 (4.5) | 94 (60.3) | 62 (39.7) |
| ≥4 years (N=141) | 136 (96.5) | 5 (3.5) | 120 (85.1) | 21 (14.9) |
| p-value^1^ | 0.8 | | <0.001 | |
| **Sex** | | | | |
| Overall (N = 504) | 481 (95.4) | 23 (4.6) | 356 (70.6) | 148 (29.4) |
| Male (N = 268) | 253 (94.4) | 15 (5.6) | 191 (71.3) | 77 (28.7) |
| Female (N = 236) | 228 (96.6) | 8 (3.4) | 165 (69.9) | 71 (30.1) |
| p-value^1^ | 0.3 | | 0.8 | |
| **Age** | | | | |
| Overall (N = 505) | 482 (95.4) | 23 (4.6) | 356 (70.5) | 149 (29.5) |
| <30 years (N = 98) | 92 (93.9) | 6 (6.1) | 75 (76.5) | 23 (23.5) |
| 30–39 years (N = 151) | 143 (94.7) | 8 (5.3) | 95 (62.9) | 56 (37.1) |
| 40–49 years (N = 130) | 124 (95.4) | 6 (4.6) | 90 (69.2) | 40 (30.8) |
| ≥50 years (N = 126) | 123 (97.6) | 3 (2.4) | 96 (76.2) | 30 (23.8) |
| p-value^1^ | 0.5 | | 0.050 | |
| **CLD subtype** | | | | |
| Overall (N = 505) | 482 (95.4) | 23 (4.6) | 356 (70.5) | 149 (29.5) |
| NAFLD/NASH/MASLD/MASH (N = 273) | 262 (96.0) | 11 (4.0) | 174 (63.7) | 99 (36.3) |
| Other (N = 232) | 220 (94.8) | 12 (5.2) | 182 (78.4) | 50 (21.6) |
| p-value^1^ | 0.7 | | <0.001 | |
| **CLD subtype** | | | | |
| Overall (N = 505) | 482 (95.4) | 23 (4.6) | 356 (70.5) | 149 (29.5) |
| Alcohol-related liver disease (N = 77) | 75 (97.4) | 2 (2.6) | 59 (76.6) | 18 (23.4) |
| Other (N = 428) | 407 (95.1) | 21 (4.9) | 297 (69.4) | 131 (30.6) |
| p-value^1^ | 0.6 | | 0.2 | |
| **CLD subtype** | | | | |
| Overall (N = 505) | 482 (95.4) | 23 (4.6) | 356 (70.5) | 149 (29.5) |
| Hepatitis (N = 144) | 137 (95.1) | 7 (4.9) | 110 (76.4) | 34 (23.6) |
| Other (N = 361) | 345 (95.6) | 16 (4.4) | 246 (68.1) | 115 (31.9) |
| p-value^1^ | 0.8 | | 0.083 | |
| **Fatigue severity** | | | | |
| Overall (N = 505) | 482 (95.4) | 23 (4.6) | 356 (70.5) | 149 (29.5) |
| MFI score ≤60 (N = 208) | 195 (93.8) | 13 (6.2) | 123 (59.1) | 85 (40.9) |
| MFI score >60 (N = 297) | 287 (96.6) | 10 (3.4) | 233 (78.5) | 64 (21.5) |
| p-value^1^ | 0.13 | | <0.001 | |
| **CLD treatment status** | | | | |
| Overall (N = 505) | 482 (95.4) | 23 (4.6) | 356 (70.5) | 149 (29.5) |
| Currently treated (N = 283) | 269 (95.1) | 14 (4.9) | 211 (74.6) | 72 (25.4) |
| Treated in the past (N = 31) | 30 (96.8) | 1 (3.2) | 25 (80.6) | 6 (19.4) |
| Never treated (N = 191) | 183 (95.8) | 8 (4.2) | 120 (62.8) | 71 (37.2) |
| p-value^1^ | >0.9 | | 0.011 | |

^1^Fisher’s exact test

CLD, chronic liver disease; MASH, metabolic dysfunction-associated steatohepatitis; MASLD, metabolic dysfunction-associated steatotic liver disease; NASH, non-alcoholic steatohepatitis; NAFLD, non-alcoholic fatty liver disease.

**Supplementary Table 8**. Mean (SD) PROMIS-29+2 T-scores by participant subgroup

|  | **Physical function^2^** | **Anxiety in past 7 days^3^** | **Depression in past 7 days^3^** | **Fatigue in past 7 days^3^** | **Sleep disturbance in past 7 days^3^** | **Ability to participate in social roles and activities^2^** | **Pain interference in past 7 days^3^** | **Cognitive function – abilities in past 7 days^2^** | **Pain intensity in past 7 days** |
| --- | --- | --- | --- | --- | --- | --- | --- | --- | --- |
| **CLD duration** | | | | | | | | | |
| Overall (N = 494) | 43.4 (7.8) | 59.5 (8.6) | 55.9 (9.3) | 55.6 (9.1) | 54.5 (6.6) | 47.5 (7.9) | 57.3 (8.6) | 43.8 (6.3) | 5.1 (2.8) |
| <1 year (N = 84) | 43.5 (7.4) | 59.1 (7.3) | 54.5 (8.5) | 55.7 (7.5) | 54.9 (6.8) | 48.2 (6.8) | 58.0 (7.9) | 44.0 (5.9) | 5.3 (2.3) |
| 1–<2 years (N = 113) | 42.3 (7.2) | 61.0 (9.7) | 57.3 (10.1) | 56.4 (9.9) | 54.7 (6.2) | 46.7 (8.0) | 58.1 (7.8) | 42.5 (6.8) | 5.4 (2.6) |
| 2–<4 years (N = 156) | 43.4 (8.1) | 57.2 (9.5) | 54.5 (9.6) | 54.0 (10.4) | 53.8 (7.0) | 48.4 (8.6) | 56.5 (9.3) | 43.9 (7.0) | 4.9 (3.0) |
| ≥4 years (N = 141) | 44.1 (8.0) | 61.1 (6.8) | 57.3 (8.6) | 56.6 (7.3) | 54.7 (6.3) | 46.8 (7.7) | 57.3 (8.7) | 44.4 (5.2) | 5.0 (3.0) |
| p-value^1^ | 0.3 | 0.003 | 0.024 | 0.2 | 0.5 | 0.2 | 0.6 | 0.2 | 0.8 |
| **Sex** | | | | | | | | | |
| Overall (N = 504) | 43.5 (7.8) | 59.4 (8.6) | 55.9 (9.3) | 55.5 (9.0) | 54.4 (6.5) | 47.6 (8.0) | 57.3 (8.6) | 43.8 (6.4) | 5.1 (2.8) |
| Male (N = 268) | 44.7 (7.9) | 58.1 (8.4) | 54.7 (9.0) | 53.7 (9.0) | 53.7 (6.3) | 48.8 (8.0) | 55.8 (8.8) | 44.0 (6.5) | 4.7 (2.9) |
| Female (N = 236) | 42.1 (7.4) | 61.0 (8.5) | 57.3 (9.5) | 57.5 (8.5) | 55.3 (6.6) | 46.2 (7.7) | 59.0 (8.0) | 43.6 (6.3) | 5.5 (2.7) |
| p-value^1^ | <0.001 | <0.001 | <0.001 | <0.001 | 0.004 | <0.001 | <0.001 | 0.4 | 0.005 |
| **Age** | | | | | | | | | |
| Overall (N = 505) | 43.5 (7.8) | 59.4 (8.6) | 55.9 (9.3) | 55.5 (9.0) | 54.4 (6.5) | 47.6 (8.0) | 57.2 (8.6) | 43.9 (6.4) | 5.1 (2.8) |
| <30 years (N = 98) | 43.5 (7.5) | 59.9 (9.1) | 56.4 (9.2) | 56.4 (8.7) | 53.9 (7.3) | 46.9 (8.0) | 58.0 (8.2) | 44.0 (6.6) | 5.6 (2.6) |
| 30–39 years (N = 151) | 45.2 (7.8) | 58.2 (9.1) | 55.3 (9.5) | 53.8 (9.9) | 54.4 (6.3) | 49.2 (8.4) | 55.5 (9.1) | 43.2 (6.4) | 4.6 (3.0) |
| 40–49 years (N = 130) | 43.7 (8.1) | 59.6 (9.0) | 55.4 (10.0) | 55.6 (9.2) | 54.2 (6.5) | 47.7 (8.1) | 57.3 (8.8) | 44.0 (6.4) | 5.0 (2.7) |
| ≥50 years (N = 126) | 41.2 (7.0) | 60.4 (6.8) | 56.8 (8.3) | 56.6 (7.6) | 55.3 (6.1) | 46.0 (7.0) | 58.6 (7.8) | 44.4 (6.2) | 5.3 (2.8) |
| p-value^1^ | <0.001 | 0.2 | 0.5 | 0.075 | 0.4 | 0.017 | 0.015 | 0.4 | 0.059 |
| **CLD subtype** | | | | | | | | | |
| Overall (N = 505) | 43.5 (7.8) | 59.4 (8.6) | 55.9 (9.3) | 55.5 (9.0) | 54.4 (6.5) | 47.6 (8.0) | 57.2 (8.6) | 43.9 (6.4) | 5.1 (2.8) |
| NAFLD/NASH/MASLD/MASH (N = 273) | 42.5 (7.7) | 58.9 (8.8) | 55.6 (9.3) | 56.0 (9.3) | 54.3 (6.9) | 47.1 (7.9) | 58.0 (8.7) | 43.7 (6.6) | 5.4 (2.8) |
| Other (N = 232) | 44.6 (7.7) | 60.0 (8.3) | 56.3 (9.4) | 54.8 (8.6) | 54.6 (6.1) | 48.1 (8.0) | 56.3 (8.4) | 44.1 (6.2) | 4.7 (2.8) |
| p-value^1^ | <0.001 | 0.3 | 0.4 | 0.13 | 0.9 | 0.049 | 0.017 | 0.7 | 0.003 |
| **CLD subtype** | | | | | | | | | |
| Overall (N = 505) | 43.5 (7.8) | 59.4 (8.6) | 55.9 (9.3) | 55.5 (9.0) | 54.4 (6.5) | 47.6 (8.0) | 57.2 (8.6) | 43.9 (6.4) | 5.1 (2.8) |
| Alcohol-related liver disease (N = 77) | 42.2 (6.6) | 60.7 (8.2) | 58.0 (8.9) | 56.8 (8.0) | 55.7 (5.2) | 45.6 (7.2) | 58.5 (6.9) | 43.0 (6.1) | 5.6 (2.4) |
| Other (N = 428) | 43.7 (7.9) | 59.2 (8.6) | 55.5 (9.3) | 55.2 (9.2) | 54.2 (6.7) | 48.0 (8.1) | 57.0 (8.9) | 44.0 (6.4) | 5.0 (2.9) |
| p-value^1^ | 0.11 | 0.7 | 0.065 | 0.2 | 0.074 | 0.047 | 0.2 | 0.2 | 0.2 |
| **CLD subtype** | | | | | | | | | |
| Overall (N = 505) | 43.5 (7.8) | 59.4 (8.6) | 55.9 (9.3) | 55.5 (9.0) | 54.4 (6.5) | 47.6 (8.0) | 57.2 (8.6) | 43.9 (6.4) | 5.1 (2.8) |
| Hepatitis (N = 144) | 45.9 (8.1) | 59.4 (8.3) | 55.1 (9.3) | 53.6 (8.8) | 54.3 (6.3) | 49.7 (8.0) | 55.1 (8.6) | 44.8 (6.1) | 4.1 (2.8) |
| Other (N = 361) | 42.5 (7.4) | 59.5 (8.7) | 56.2 (9.3) | 56.2 (9.0) | 54.5 (6.6) | 46.8 (7.8) | 58.1 (8.4) | 43.5 (6.5) | 5.5 (2.7) |
| p-value^1^ | <0.001 | 0.9 | 0.3 | 0.004 | 0.4 | <0.001 | <0.001 | 0.13 | <0.001 |
| **Fatigue severity** | | | | | | | | | |
| Overall (N = 505) | 43.5 (7.8) | 59.4 (8.6) | 55.9 (9.3) | 55.5 (9.0) | 54.4 (6.5) | 47.6 (8.0) | 57.2 (8.6) | 43.9 (6.4) | 5.1 (2.8) |
| MFI score ≤60 (N = 144) | 47.9 (7.3) | 54.5 (7.6) | 50.3 (8.0) | 49.6 (8.2) | 51.5 (6.1) | 52.8 (7.0) | 52.6 (7.8) | 45.6 (7.0) | 3.8 (2.8) |
| MFI score >60 (N = 361) | 40.4 (6.5) | 62.9 (7.4) | 59.8 (8.1) | 59.6 (7.1) | 56.5 (6.1) | 43.9 (6.5) | 60.5 (7.5) | 42.7 (5.6) | 6.0 (2.5) |
| p-value^1^ | <0.001 | <0.001 | <0.001 | <0.001 | <0.001 | <0.001 | <0.001 | <0.001 | <0.001 |
| **CLD treatment status** | | | | | | | | | |
| Overall (N = 505) | 43.5 (7.8) | 59.4 (8.6) | 55.9 (9.3) | 55.5 (9.0) | 54.4 (6.5) | 47.6 (8.0) | 57.2 (8.6) | 43.9 (6.4) | 5.1 (2.8) |
| Currently treated (N = 283) | 43.3 (7.5) | 59.8 (7.3) | 56.0 (8.6) | 55.8 (8.4) | 54.4 (6.5) | 47.4 (7.3) | 57.6 (8.4) | 44.3 (5.7) | 5.1 (2.8) |
| Treated in the past (N = 31) | 43.7 (8.3) | 58.3 (9.6) | 55.0 (9.2) | 56.6 (8.8) | 54.8 (7.3) | 48.4 (9.7) | 57.9 (9.3) | 45.5 (7.5) | 5.0 (2.6) |
| Never treated (N = 191) | 43.7 (8.0) | 59.1 (10.1) | 55.9 (10.3) | 54.7 (9.9) | 54.5 (6.5) | 47.7 (8.7) | 56.6 (8.8) | 42.9 (7.0) | 5.1 (2.9) |
| p-value^1^ | >0.9 | 0.8 | 0.9 | 0.5 | >0.9 | 0.5 | 0.8 | 0.11 | 0.8 |

^1^Kruskal-Wallis rank sum test

^2^The higher the score, the lower the impact on the participant

^3^The higher the score, the greater the impact on the participant

CLD, chronic liver disease; MASH, metabolic dysfunction-associated steatohepatitis; MASLD, metabolic dysfunction-associated steatotic liver disease; MFI, Multidimensional Fatigue Inventory; NASH, non-alcoholic steatohepatitis; NAFLD, non-alcoholic fatty liver disease; PROMIS-29+2, Patient-Reported Outcomes Measurement Information System.

**Supplementary Table 9**. WPAI GH scores by participant subgroup

|  | **Percentage work time missed due to fatigue (on those currently employed)** | **Percentage impairment while working due to fatigue (on those who have reported hours actually worked** | **Percentage overall work impairment due to fatigue (on those who have reported hours actually worked)** | **Percentage class time missed due to fatigue (on those currently attending classes in an academic setting)** | **Percentage impairment in the classroom due to fatigue (on those who have reported hours of school or class attended)** | **Percentage overall classroom impairment due to fatigue (on those who have reported hours of school or class attended)** | **Percentage activity impairment due to fatigue** |
| --- | --- | --- | --- | --- | --- | --- | --- |
| **CLD duration** | | | | | | | |
| Overall (N = 494) | 16.1 (19.3) | 50.2 (26.9) | 55.0 (28.4) | 27.3 (23.0) | 56.6 (25.2) | 65.7 (26.5) | 53.3 (26.3) |
| <1 year (N = 84) | 10.9 (11.2) | 47.4 (22.8) | 52.2 (23.4) | 24.2 (16.9) | 54.0 (22.9) | 63.7 (25.2) | 52.4 (23.0) |
| 1–<2 years (N = 113) | 20.7 (22.0) | 57.9 (26.0) | 62.6 (27.2) | 38.1 (27.4) | 61.3 (18.1) | 73.8 (10.3) | 58.1 (25.8) |
| 2–<4 years (N = 156) | 14.4 (16.3) | 44.4 (27.9) | 49.8 (29.8) | 15.3 (26.0) | 46.0 (40.4) | 48.7 (43.6) | 48.8 (27.9) |
| ≥4 years (N = 141) | 16.9 (22.4) | 51.6 (27.1) | 55.9 (29.2) | 29.4 (27.7) | 70.0 (25.8) | 77.8 (26.1) | 55.1 (26.2) |
| p-value^1^ | 0.13 | 0.010 | 0.021 | 0.3 | 0.6 | 0.5 | 0.039 |
| **Sex** | | | | | | | |
| Overall (N = 504) | 15.9 (19.1) | 49.8 (26.9) | 54.6 (28.5) | 26.5 (23.1) | 55.8 (25.3) | 64.6 (26.9) | 53.1 (26.4) |
| Male (N = 268) | 14.8 (18.9) | 45.3 (26.5) | 50.2 (28.7) | 27.5 (29.4) | 54.6 (29.9) | 61.5 (33.1) | 48.3 (26.9) |
| Female (N = 236) | 17.8 (19.5) | 57.9 (25.9) | 62.4 (26.6) | 25.8 (18.3) | 56.5 (22.5) | 66.6 (22.6) | 58.6 (24.7) |
| p-value^1^ | 0.034 | <0.001 | <0.001 | 0.8 | >0.9 | >0.9 | <0.001 |
| **Age** | | | | | | | |
| Overall (N = 505) | 15.8 (19.1) | 49.7 (27.0) | 54.5 (28.6) | 26.5 (23.1) | 55.8 (25.3) | 64.6 (26.9) | 53.0 (26.4) |
| <30 years (N = 98) | 22.3 (22.2) | 52.1 (28.2) | 58.0 (30.0) | 25.1 (23.3) | 54.5 (26.5) | 62.2 (29.0) | 55.6 (26.9) |
| 30–39 years (N = 151) | 12.6 (16.3) | 49.9 (27.0) | 53.9 (28.0) | 38.8 (30.7) | 70.0 (25.8) | 79.7 (27.0) | 50.6 (26.6) |
| 40–49 years (N = 130) | 15.4 (20.3) | 47.9 (26.9) | 52.4 (28.9) | 26.2 (3.4) | 60.0 (14.1) | 70.7 (9.1) | 51.6 (27.8) |
| ≥50 years (N = 126) | 15.9 (17.1) | 49.8 (26.2) | 55.4 (27.8) | 23.0 (23.4) | 48.0 (23.9) | 60.3 (21.7) | 55.3 (24.2) |
| p-value^1^ | 0.020 | 0.8 | 0.6 | 0.8 | 0.7 | 0.3 | 0.4 |
| **CLD subtype** | | | | | | | |
| Overall (N = 505) | 15.8 (19.1) | 49.7 (27.0) | 54.5 (28.6) | 26.5 (23.1) | 55.8 (25.3) | 64.6 (26.9) | 53.0 (26.4) |
| NAFLD/NASH/MASLD/MASH (N = 273) | 14.9 (17.3) | 50.6 (26.3) | 55.6 (27.5) | 31.4 (18.9) | 56.7 (25.4) | 69.0 (25.5) | 54.5 (27.0) |
| Other (N = 232) | 16.7 (20.8) | 48.9 (27.7) | 53.4 (29.6) | 20.9 (26.6) | 54.7 (25.9) | 59.3 (28.3) | 51.3 (25.7) |
| p-value^1^ | >0.9 | 0.5 | 0.6 | 0.078 | 0.7 | 0.2 | 0.087 |
| **CLD subtype** | | | | | | | |
| Overall (N = 505) | 15.8 (19.1) | 49.7 (27.0) | 54.5 (28.6) | 26.5 (23.1) | 55.8 (25.3) | 64.6 (26.9) | 53.0 (26.4) |
| Alcohol-related liver disease (N = 77) | 21.7 (22.8) | 58.9 (25.4) | 63.6 (26.3) | 36.5 (30.9) | 66.7 (23.4) | 73.3 (24.0) | 59.6 (25.0) |
| Other (N = 428) | 14.5 (17.9) | 47.6 (26.9) | 52.4 (28.7) | 23.9 (20.6) | 53.3 (25.4) | 62.6 (27.5) | 51.8 (26.5) |
| p-value^1^ | 0.017 | 0.006 | 0.007 | 0.4 | 0.3 | 0.3 | 0.029 |
| **CLD subtype** | | | | | | | |
| Overall (N = 505) | 15.8 (19.1) | 49.7 (27.0) | 54.5 (28.6) | 26.5 (23.1) | 55.8 (25.3) | 64.6 (26.9) | 53.0 (26.4) |
| Hepatitis (N = 144) | 13.4 (18.7) | 43.2 (26.1) | 47.7 (28.6) | 8.3 (14.4) | 47.1 (31.5) | 49.2 (33.7) | 46.3 (25.2) |
| Other (N = 361) | 16.9 (19.2) | 52.7 (26.9) | 57.6 (28.1) | 31.2 (22.8) | 58.1 (23.5) | 68.7 (23.8) | 55.7 (26.4) |
| p-value^1^ | 0.027 | 0.003 | 0.005 | 0.015 | 0.5 | 0.2 | <0.001 |
| **Fatigue severity** | | | | | | | |
| Overall (N = 505) | 15.8 (19.1) | 49.7 (27.0) | 54.5 (28.6) | 26.5 (23.1) | 55.8 (25.3) | 64.6 (26.9) | 53.0 (26.4) |
| MFI score ≤60 (N = 208) | 8.4 (16.2) | 34.7 (25.3) | 38.0 (27.0) | 13.9 (18.2) | 46.9 (30.4) | 52.9 (31.6) | 35.9 (25.6) |
| MFI score >60 (N = 297) | 22.0 (19.2) | 62.1 (21.5) | 68.1 (21.9) | 34.2 (22.7) | 61.5 (20.1) | 72.1 (20.8) | 65.0 (19.5) |
| p-value^1^ | <0.001 | <0.001 | <0.001 | 0.011 | 0.2 | 0.053 | <0.001 |
| **CLD treatment status** | | | | | | | |
| Overall (N = 505) | 15.8 (19.1) | 49.7 (27.0) | 54.5 (28.6) | 26.5 (23.1) | 55.8 (25.3) | 64.6 (26.9) | 53.0 (26.4) |
| Currently treated (N = 283) | 15.7 (18.5) | 48.7 (24.9) | 54.2 (26.8) | 17.4 (19.6) | 47.7 (22.8) | 56.3 (23.8) | 53.3 (23.7) |
| Treated in the past (N = 31) | 13.7 (17.3) | 44.3 (26.2) | 48.9 (28.9) | 25.0 (35.4) | 25.0 (35.4) | 37.5 (53.0) | 42.6 (27.7) |
| Never treated (N = 191) | 16.3 (20.3) | 52.1 (29.8) | 55.9 (31.0) | 32.8 (23.5) | 65.0 (22.6) | 73.5 (23.8) | 54.2 (29.6) |
| p-value^1^ | 0.8 | 0.3 | 0.5 | 0.3 | 0.023 | 0.027 | 0.046 |

CLD, chronic liver disease; MASH, metabolic dysfunction-associated steatohepatitis; MASLD, metabolic dysfunction-associated steatotic liver disease; MFI, Multidimensional Fatigue Inventory; NASH, non-alcoholic steatohepatitis; NAFLD, non-alcoholic fatty liver disease; WPAI:SHP, Work Productivity and Activity Impairment – Specific Health Problem.

**Supplementary Table 10**. MFI scores by participant subgroup

|  | **General fatigue (items 1, 5, 12, 16)** | **Physical fatigue (items 2, 8, 14, 20)** | **Reduced activity (items 3, 6, 10, 17)** | **Reduced motivation (items 4, 9, 15, 18)** | **Mental fatigue (items 7, 11, 13, 19)** | **Total MFI score (all items)** |
| --- | --- | --- | --- | --- | --- | --- |
| **CLD duration** | | | | | | |
| Overall (N = 494) | 13.9 (3.4) | 13.4 (3.6) | 12.3 (3.6) | 11.2 (3.3) | 12.5 (3.7) | 63.3 (14.6) |
| <1 year (N = 84) | 13.8 (3.2) | 12.8 (3.9) | 11.9 (3.9) | 10.7 (3.5) | 11.7 (3.8) | 60.9 (15.8) |
| 1–<2 years (N = 113) | 14.3 (3.2) | 13.7 (3.4) | 12.6 (3.5) | 11.9 (3.6) | 13.1 (3.8) | 65.7 (15.0) |
| 2–<4 years (N = 156) | 13.2 (3.8) | 13.2 (3.6) | 12.2 (3.6) | 10.9 (3.6) | 11.7 (3.9) | 61.3 (15.6) |
| ≥4 years (N = 141) | 14.5 (3.1) | 13.8 (3.4) | 12.4 (3.5) | 11.0 (2.6) | 13.3 (3.1) | 65.0 (11.6) |
| p-value^1^ | 0.040 | 0.2 | 0.5 | 0.077 | <0.001 | 0.035 |
| **Sex** | | | | | | |
| Overall (N = 504) | 13.9 (3.4) | 13.4 (3.6) | 12.3 (3.6) | 11.1 (3.3) | 12.4 (3.7) | 63.2 (14.6) |
| Male (N = 268) | 13.4 (3.3) | 12.8 (3.4) | 11.8 (3.5) | 10.8 (3.2) | 12.0 (3.6) | 60.8 (14.0) |
| Female (N = 236) | 14.5 (3.4) | 14.1 (3.6) | 12.8 (3.6) | 11.5 (3.5) | 13.0 (3.8) | 65.9 (14.8) |
| p-value^1^ | <0.001 | <0.001 | <0.001 | 0.007 | 0.004 | <0.001 |
| **Age** | | | | | | |
| Overall (N = 505) | 13.9 (3.4) | 13.4 (3.6) | 12.3 (3.6) | 11.1 (3.4) | 12.4 (3.7) | 63.1 (14.6) |
| <30 years (N = 98) | 14.1 (3.6) | 13.4 (4.0) | 13.1 (4.0) | 11.9 (3.6) | 12.8 (3.9) | 65.4 (17.0) |
| 30–39 years (N = 151) | 13.2 (3.4) | 12.9 (3.6) | 11.8 (3.4) | 11.0 (3.3) | 12.3 (3.5) | 61.2 (13.9) |
| 40–49 years (N = 130) | 13.7 (3.4) | 13.0 (3.5) | 11.9 (3.7) | 10.7 (3.5) | 12.3 (3.9) | 61.6 (14.8) |
| ≥50 years (N = 126) | 14.8 (3.0) | 14.3 (3.3) | 12.6 (3.3) | 11.1 (2.9) | 12.5 (3.7) | 65.3 (12.8) |
| p-value^1^ | 0.002 | 0.003 | 0.021 | 0.074 | 0.7 | 0.021 |
| **CLD subtype** | | | | | | |
| Overall (N = 505) | 13.9 (3.4) | 13.4 (3.6) | 12.3 (3.6) | 11.1 (3.4) | 12.4 (3.7) | 63.1 (14.6) |
| NAFLD/NASH/MASLD/MASH (N = 273) | 13.6 (3.4) | 13.3 (3.5) | 12.3 (3.4) | 11.0 (3.2) | 12.1 (3.7) | 62.2 (14.0) |
| Other (N = 232) | 14.3 (3.3) | 13.5 (3.7) | 12.3 (3.8) | 11.3 (3.5) | 12.9 (3.7) | 64.2 (15.3) |
| p-value^1^ | 0.068 | 0.5 | 0.7 | 0.3 | 0.020 | 0.3 |
| **CLD subtype** | | | | | | |
| Overall (N = 505) | 13.9 (3.4) | 13.4 (3.6) | 12.3 (3.6) | 11.1 (3.4) | 12.4 (3.7) | 63.1 (14.6) |
| Alcohol-related liver disease (N = 77) | 14.9 (3.4) | 14.4 (4.0) | 13.2 (4.2) | 12.0 (3.7) | 13.4 (4.2) | 67.9 (16.9) |
| Other (N = 428) | 13.7 (3.4) | 13.2 (3.5) | 12.1 (3.4) | 11.0 (3.3) | 12.3 (3.6) | 62.2 (14.0) |
| p-value^1^ | 0.008 | 0.010 | 0.044 | 0.027 | 0.024 | 0.010 |
| **CLD subtype** | | | | | | |
| Overall (N = 505) | 13.9 (3.4) | 13.4 (3.6) | 12.3 (3.6) | 11.1 (3.4) | 12.4 (3.7) | 63.1 (14.6) |
| Hepatitis (N = 144) | 13.6 (3.3) | 12.9 (3.5) | 11.6 (3.3) | 10.7 (3.3) | 12.4 (3.2) | 61.2 (13.6) |
| Other (N = 361) | 14.0 (3.4) | 13.6 (3.6) | 12.6 (3.6) | 11.3 (3.4) | 12.4 (3.9) | 63.9 (14.9) |
| p-value^1^ | 0.2 | 0.067 | 0.006 | 0.11 | >0.9 | 0.045 |
| **Fatigue severity** | | | | | | |
| Overall (N = 505) | 13.9 (3.4) | 13.4 (3.6) | 12.3 (3.6) | 11.1 (3.4) | 12.4 (3.7) | 63.1 (14.6) |
| MFI score ≤60 (N = 208) | 11.3 (2.7) | 10.3 (2.3) | 9.5 (2.4) | 8.5 (2.6) | 9.6 (2.7) | 49.3 (8.4) |
| MFI score >60 (N = 297) | 15.8 (2.5) | 15.5 (2.6) | 14.2 (3.0) | 12.9 (2.6) | 14.4 (3.0) | 72.8 (9.2) |
| p-value^1^ | <0.001 | <0.001 | <0.001 | <0.001 | <0.001 | <0.001 |
| **CLD treatment status** | | | | | | |
| Overall (N = 505) | 13.9 (3.4) | 13.4 (3.6) | 12.3 (3.6) | 11.1 (3.4) | 12.4 (3.7) | 63.1 (14.6) |
| Currently treated (N = 283) | 14.2 (3.1) | 13.7 (3.5) | 12.4 (3.3) | 11.1 (3.1) | 12.7 (3.3) | 64.1 (12.9) |
| Treated in the past (N = 31) | 13.4 (2.9) | 12.3 (3.9) | 11.6 (4.2) | 10.5 (3.2) | 11.3 (3.7) | 59.1 (14.9) |
| Never treated (N = 191) | 13.6 (3.8) | 13.2 (3.7) | 12.1 (3.9) | 11.3 (3.7) | 12.2 (4.2) | 62.3 (16.7) |
| p-value^1^ | 0.2 | 0.060 | 0.2 | 0.5 | 0.034 | 0.055 |

CLD, chronic liver disease; MASH, metabolic dysfunction-associated steatohepatitis; MASLD, metabolic dysfunction-associated steatotic liver disease; MFI, Multidimensional Fatigue Inventory; NASH, non-alcoholic steatohepatitis; NAFLD, non-alcoholic fatty liver disease.
